# Suicide trends in Portugal from 2002–2023 - a time-series analysis pondering data structure and fluctuations of undetermined intent and accidental deaths

**DOI:** 10.64898/2026.07.16.26358214

**Authors:** Edgar Mesquita, Virgínia da Conceição, Ricardo Gusmão

**Affiliations:** EPIUnit, Institute of Public Health of the University of Porto, Portugal; Laboratory for Integrative and Translational Research in Population Health (ITR), Porto, Portugal; Department of Clinical Neurosciences and Mental Health, Faculty of Medicine, University of Porto, Portugal

**Keywords:** Suicide, Misclassification, Mortality trends, Joinpoint regression, Undetermined intent, Surveillance

## Abstract

**Purpose:** Suicide mortality is underestimated due to misclassification under undetermined and accidental deaths. This study examined national trends in suicide and related external causes of death in Portugal from 2002 to 2023, by sex and age group, assessing potential shifts suggesting masked suicide and quantifying the relationship between undetermined, suicide, and accident death rates through ratio indices.

**Methods:** Using official mortality data from Portugal’s Statistics Institute (INE) for 2002– 2023, we calculated age-standardised (SDR) and age-specific death rates (ASDR) for suicide (X60–X84), undetermined intent deaths (Y10–Y34), and unintentional deaths (V01–X59), disaggregated by sex and four age groups (15–24, 25–44, 45–64, ≥65). We estimated undetermined-to-suicide (UnD:Suic) and undetermined-to-accidents (UnD:Accs) rate ratios for SDRs and ASDRs. Trends were analysed using joinpoint regression (APC/AAPC) and structural breakpoint analysis (Chow test, BIC).

**Results:** Suicide SDRs declined across the period for males (AAPC: −2.25%) and females (AAPC: −1.32%), with the sharpest reductions among males aged 25–44 (AAPC: −2.56%) and females aged 65+ (AAPC: −2.44%). Deaths of undetermined intent rose steeply from 2002 to 2005–2006 and declined thereafter. Unintentional deaths declined in most age groups, except females aged 65+ (AAPC: +1.41%). Both ratio series peaked around 2005– 2009, declined progressively through the 2010s, and reached their lowest values in 2021– 2022. Age-specific analyses revealed a significant and sustained increase in both ratios among females aged 45–64. Structural breakpoints clustered around 2004, 2013–2015, and 2019–2020.

**Conclusion:** Suicide mortality declined in Portugal from 2002 to 2023, but divergent trends in undetermined and accidental deaths across sex and age subgroups highlight ongoing misclassification. Age- and sex-specific ratio analyses identify the population subgroups where misclassification is most concentrated, providing a foundation for future imputation-based estimates of probable suicide burden.

## 1. Background

Suicide is a public mental health first-order priority in Portugal, with a mortality burden higher than other Southern European countries and approaching the European Union average [1,2].

The regular update of national suicide trends is necessary to enhance accurate recognition of evolving mental health care needs and the establishment of priorities to enable and adjust suicide prevention interventions [3].

However, suicide trend assessment is complicated by the extent of misclassification under undetermined intent deaths, accidental deaths, and ill-defined causes [4-6]. Psychological autopsy research further documents the prevalence of concealment of suicidal intent, both conscious and unconscious, which complicates the attribution of intent at death certification [7]. The WHO has recognised certain death categories as ‘garbage codes’ and redistributes them to suicide when calculating country and global mortality rates [8].

Suicide concealment reaches a serious magnitude in Portugal, with undetermined-to-suicide ratios that have historically surpassed 2:1 [9-13].

In previous research spanning 1913 to 2018, we demonstrated systematic misclassification and errors in the Portuguese death registry and identified the improved accuracy of suicide and undetermined death certification associated with a verbal autopsy procedure implemented in 2002–2003, coinciding with the adoption of ICD-10 coding [2]. Since the beginning of the 21st century, further changes within the statistical system and at broader social and economic levels have likely affected registry quality and mortality trends [2].

At least five significant procedural changes in Portugal’s external death registration can be identified during the study period. First, ICD-10 was introduced in 2002. Second, the Directorate General of Health conducted a verbal inquiry with medical doctors who issued death certificates between 2001 and 2004 [2]. Third, Portugal failed to report data for 2004, 2005, and 2006 to the WHO Mortality Database [14]. Fourth, death certification was digitalised between 2013 and 2015 through the Death Certificate Information System (SICO) [15,16]. Fifth, during the COVID-19 pandemic in 2020–2021, Portugal again failed to report data to the WHO Mortality Database [14]. In parallel, major socioeconomic events - the 2010–2014 financial crisis and austerity period, and the COVID-19 pandemic from 2020 - may have independently affected both actual suicide rates and their classification [17-21].

The impact of the SICO digitalisation on suicide trend classification has not been previously examined. A national suicide prevention strategy was designed for 2013–2017 but was never systematically implemented [22].

The gaps in interpreting suicide data produced by Statistics Portugal undermine the knowledge of suicide trends: the examination of national suicide trends should consider other external death causes, especially where there is erratic fluctuation in data consistency and a higher proportion of undetermined deaths and accidents.

### 1.1 Purpose of the study

Our primary aim is to characterise the distribution and trends in registered suicide, undetermined intent, and unintentional mortality in Portugal from 2002 to 2023, by sex and age. Our secondary aim is to quantify the temporal relationship between these three categories through undetermined-to-suicide (UnD:Suic) and undetermined-to-accidents (UnD:Accs) rate ratios, disaggregated by sex and age group, to identify the population subgroups where suicide concealment is most likely occurring and to prepare the evidential basis for future imputation-based estimates of probable suicide burden.

We expect the suicide accuracy registry (i) to improve during 2010–2014 through the suicide awareness-raising impact on coders; (ii) to continue to improve from 2014 through 2016, secondarily to the digitalisation of the vital death registry; (iii) to improve again from 2020– 2021 because of death scrutiny to accurately report COVID deaths. We also assume (iv) a fade-out effect from 2004–2009 and from 2017 to 2019, with undetermined and accidental deaths increasing, corresponding to the accrued concealment of suicide; and (v) that undetermined intent deaths function as a reservoir for both misclassified suicides and misclassified accidents.

## 2 Methods

### 2.1 Data collection

We obtained yearly population and mortality data from Portugal’s Statistics Institute (INE) [23] from 2002 to 2023. These official datasets include annual end-of-year population and mortality records classified by residence, sex, and five-year age groups, coded according to ICD-10 Chapter 20 [24] and aggregated by intent according to the European Shortlist for Causes of Death [25]: registered suicide and intentionally self-inflicted injuries (X60–X84), registered undetermined intent deaths (Y10–Y34), and registered unintentional deaths (V01– X59). Homicides were excluded because, under the Portuguese legal system, they are systematically investigated through legal inquiry, making misclassification into this category unlikely [26]. The study period begins in 2002, when ICD-10 was adopted in Portugal, avoiding a significant data breakpoint due to the ICD-9/ICD-10 transition [2].

### 2.2 Data analysis

Population figures were derived from intercensal estimates; mid-year estimates (1 July) were used to calculate age-specific death rates (ASDRs) within age groups 15–24, 25–44, 45–64, and ≥65. Age-standardised death rates (SDRs) were computed by the direct method [27] using the 2013 European Standard Population [28], for suicide, undetermined intent deaths, and unintentional deaths, separately for males and females. Annual absolute numbers and male-to-female ratios were also presented for each category.

We then estimated undetermined-to-suicide rate ratios (UnD:Suic) and undetermined-to-accidents rate ratios (UnD:Accs) by dividing undetermined intent SDRs and ASDRs by the corresponding suicide or accident SDRs and ASDRs, respectively. These ratios operationalise the magnitude of potential misclassification and allow temporal and subgroup comparisons. The UnD:Suic ratio and the UnD:Accs ratio both describe the relative weight of undetermined intent deaths against different reference categories, revealing qualitatively different information. The UnD:Suic ratio captures the extent to which undetermined deaths may be absorbing misclassified suicides; the UnD:Accs ratio captures the extent to which they may be absorbing misclassified accidents. Together, they allow an assessment of whether undetermined deaths function primarily as a reservoir for concealed suicides, for misclassified accidents, or for both simultaneously.

Joinpoint regression analysis was applied to SDRs and ASDRs of all three mortality categories and to both ratio series, separately for males and females and by age group, estimating the annual percent change (APC) for each segment and the average annual percent change (AAPC) for the overall 2002–2023 period [29].

Structural breakpoint analysis was conducted for each SDR and ASDR time series using the Bayesian Information Criterion (BIC) to identify breakpoints, which were verified by the Chow test (p<0.01) using the strucchange package in R [2,30]. All statistical analyses were conducted using R software [31] and Joinpoint Version 5.3.0.0 (National Cancer Institute) [32].

### 2.3 Ethical considerations

Mortality and population data from Portugal are publicly available, aggregated at population level, anonymised, and consist of routinely collected administrative data from INE [23]. Ethics approval was not required.

## 3. Results

Over the 22-year period, 99,639 deaths were registered (4.12% of all 2,416,403 deaths): 22,828 by suicide (22.9%), 17,155 by undetermined intent (17.2%), and 59,656 as unintentional or accidental (56.9%). Males accounted for 67.4% of all violent deaths. Deaths concentrated in the ≥65 age group (54.3%), with young people aged 15–24 least affected (5.1%) (Table 1).

**Table 1.** Distribution of external causes of death in Portugal by sex and age group, 2002– 2023 (INE)

|  |  | Total |  | Males |  | Females |  | % of deaths:<br>Males |
| --- | --- | --- | --- | --- | --- | --- | --- | --- |
|  |  | n | % | n | % | n | % |  |
| Suicide | **Total** | **22 828** |  | **17 491** |  | **5 337** |  | **76.6** |
|  | **15-24** | 849 | 3.7 | 669 | 3.8 | 180 | 3.4 | 78.8 |
|  | **25-44** | 4 642 | 20.3 | 3 695 | 21.1 | 947 | 17.7 | 79.6 |
|  | **45-64** | 7 769 | 34.0 | 5 866 | 33.5 | 1 903 | 35.7 | 75.5 |
|  | **>65** | 9 516 | 41.7 | 7 226 | 41.3 | 2 290 | 42.9 | 75.9 |
| Undetermined deaths | **Total** | **17 155** |  | **10 270** |  | **6 883** |  | **59.9** |
|  | **15-24** | 449 | 2.6 | 355 | 3.5 | 94 | 1.4 | 79.1 |
|  | **25-44** | 1 940 | 11.3 | 1 579 | 15.4 | 361 | 5.2 | 81.4 |
|  | **45-64** | 3 427 | 20.0 | 2 708 | 26.4 | 719 | 10.4 | 79.0 |
|  | **>65** | 11 147 | 65.0 | 5 497 | 53.5 | 5 650 | 82.1 | 49.3 |
| All unintentional deaths | **Total** | **59 656** |  | **39 422** |  | **20 272** |  | **66.1** |
|  | **15-24** | 3 795 | 6.4 | 3 143 | 8.0 | 652 | 3.2 | 82.8 |
|  | **25-44** | 9 415 | 15.8 | 8 160 | 20.7 | 1 255 | 6.2 | 86.7 |
|  | **45-64** | 11 945 | 20.0 | 9 688 | 24.6 | 2 257 | 11.1 | 81.1 |
|  | **>65** | 33 395 | 56.0 | 17 697 | 44.9 | 15 698 | 77.4 | 53.0 |
| All Violent deaths | **Total** | **99 639** |  | **67 183** |  | **32 492** |  | **67,4** |
|  | **15-24** | 5 093 | 5,1 | 4 167 | 6,2 | 926 | 2,8 | 81,8 |
|  | **25-44** | 15 997 | 16,1 | 13 434 | 20,0 | 2 563 | 7,9 | 84,0 |
|  | **45-64** | 23 141 | 23,2 | 18 262 | 27,2 | 4 879 | 15,0 | 78,9 |
|  | **>65** | 54 058 | 54,3 | 30 420 | 45,3 | 23 638 | 72,8 | 56,3 |

### 3.1 Suicide, undetermined intent, and unintentional official death trends

Visually, an inverse relation is apparent between undetermined intent death rates, both with accidental death rates and with suicide rates, for both sexes (Figure 1).

**Figure 1.**
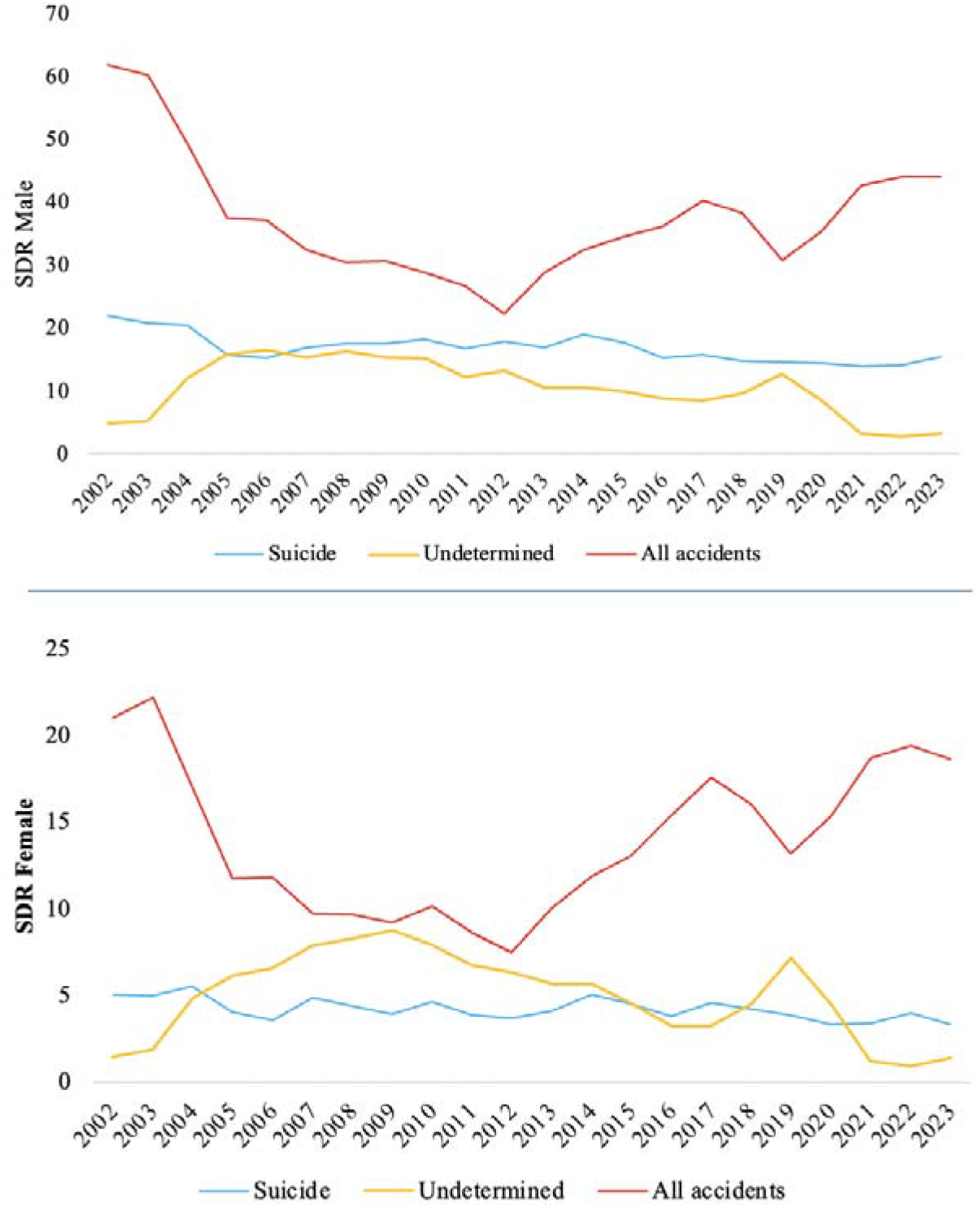
SDRs of suicide, undetermined intent, and accidents in Portugal by sex, 2002–2023.

We observed a decline in suicide, undetermined intent, and unintentional mortality across 2002–2023 for both males and females, similar in AAPC for suicide and undetermined intent, but steeper for males; for accidents, the reduction was substantially larger for males than for females (Figure 1). The general reduction tendency is opposed by two segments of increase for both sexes: a steep rise from 2002 to 2005/2006 for undetermined SDRs, and a sustained increase from approximately 2008/2010 onwards for accident SDRs.

Overall, breakpoints are similar in males and females. For males, the key breakpoint is 2004: up to 2004, accident SDRs decreased and undetermined SDRs increased; thereafter, they reversed. For females, the breakpoints are similar, with no significant changes in segment slopes (Figure 2, supplementary Tables S12 and S13).

**Figure 2.**
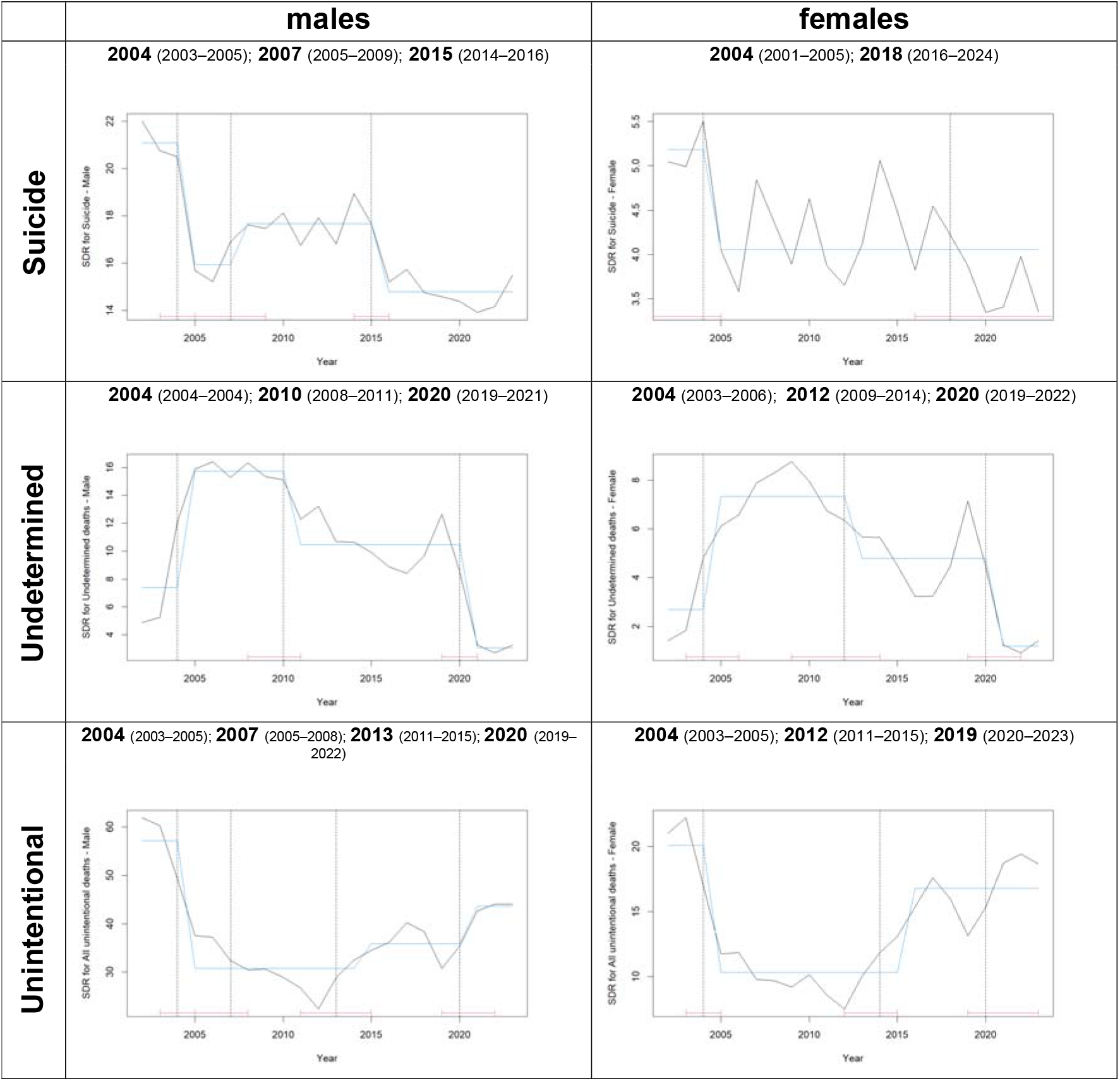
Structural breakpoints in age-standardised death rates (SDRs) for suicide, undetermined deaths, and unintentional deaths in Portugal by sex, 2002–2023.

Among males, suicide SDRs declined significantly in all age groups except 15–24, with the largest decline in the 25–44 group. For undetermined deaths, reductions were observed across all male age groups, with the sharpest statistically significant decline in the 65+ group. Accident SDRs decreased significantly in all male age groups below 65, most markedly in 15–24 and 25–44.

Among females, suicide SDRs decreased in all age groups, most significantly in the 65+ group. Undetermined deaths increased significantly in the 45–64 group, while decreasing in the 65+ group. Accident SDRs decreased in all female age groups except 65+ (AAPC: +1.41%; 95% CI: 0.15–2.68) (Table 2).

**Table 2.** Average annual percentage change (AAPC) in age-standardised death rates (ASDRs) and ratios by age group and sex, Portugal, 2002–2023.

|  | Males |  |  |  | Females |  |  |  |
| --- | --- | --- | --- | --- | --- | --- | --- | --- |
| AAPC<br>(95% CI) | 15–24 | 25–44 | 45–64 | ≥65 | 15–24 | 25–44 | 45–64 | ≥65 |
| **Suicide** | -0.21<br>(-2.28; 2.02) | -2.56*<br>(-3.60; -1.69) | -0.66<br>(-1.40; 0.03) | -2.25*<br>(-2.87; -1.58) | 2.57*<br>(0.07; 5.26) | -1.49<br>(-3.16; 0.31) | -0.55<br>(-2.00; 0.98) | -2.44*<br>(-3.89; -0.66) |
| **Undetermined deaths** | -2.74<br>(-7.57; 1.30) | -1.12<br>(-3.90; 1.88) | -0.38<br>(-2.33; 1.46) | -3.75<br>(-7.28; 0.07) | -6.11<br>(-14.06; 2.81) | 0.23<br>(-5.88; 4.90) | 6.23*<br>(3.96; 9.01) | -3.05<br>(-7.02; 1.23) |
| **All unintentional deaths** | -5.75*<br>(-6.92; -4.63) | -4.60*<br>(-5.78; -3.33) | -2.00*<br>(-3.02; -0.84) | 1.14<br>(-0.16; 2.60) | -5.69*<br>(-8.86; -2.67) | -5.15*<br>(-7.07; -3.07) | -3.34*<br>(-4.49; -1.93) | 1.41*<br>(0.15; 2.68) |
| **Ratio UnD:Suic** | -1.58<br>(-5.07; 1.92) | 0.39<br>(-2.32; 3.76) | -0.11<br>(-2.92; 2.95) | -1.76<br>(-5.46; 2.14) | -8.46<br>(-16.32; 0.56) | 1.12<br>(-3.17; 6.10) | 7.73*<br>(5.03; 11.48) | -0.23<br>(-5.06; 4.91) |
| **Ratio UnD:Accs** | 4.66*<br>(1.92; 8.27) | 2.67<br>(-0.18; 5.61) | 1.07<br>(-1.43; 3.56) | -4.87<br>(-10.34; 1.32) | -0.03<br>(-8.45; 9.37) | 5.23*<br>(0.28; 10.71) | 10.77*<br>(7.83; 13.74) | -1.76<br>(-7.22; 4.92) |

\*Statistically significant (95% CI excludes zero).

### 3.2 Ratios of undetermined intent to suicide (UnD:Suic) and undetermined intent to accidents (UnD:Accs)

At the SDR level, both ratios shared a strikingly similar temporal pattern. Both were low and stable in 2002–2003 (UnD:Suic total: 0.24 and 0.29; UnD:Accs total: 0.07 and 0.08), then increased sharply in 2004–2006 following the suspension of the verbal autopsy procedure, peaking between 2006 and 2009 when undetermined deaths reached their highest levels relative to both suicide and accidents. The UnD:Suic ratio exceeded 1.0 in females from 2005 to 2010 (peak: 2.25 in 2009) and approached but did not consistently exceed 1.0 in males (peak: 1.08 in 2006). Both ratios then declined progressively, with a transient increase in 2019 coinciding with a spike in undetermined deaths, before falling sharply in 2021–2022 to their lowest values across the entire study period (UnD:Suic total 2022: 0.21; UnD:Accs total 2022: 0.06). A partial recovery in 2023 suggests that misclassification resumed as pandemic-era scrutiny relaxed (Figure 3, supplementary Tables S4–S5).

**Figure 3.**
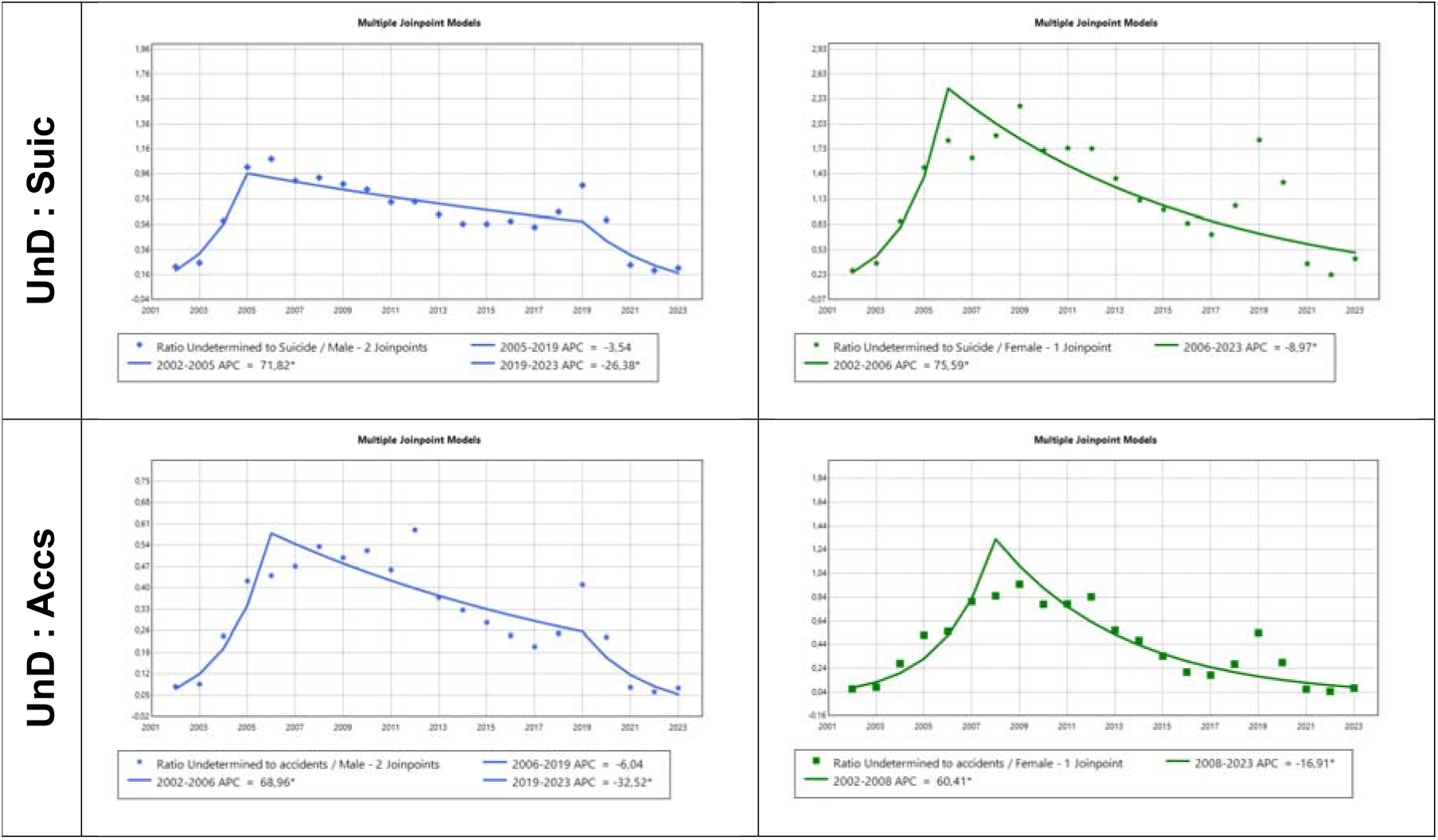
Trends in age-standardised death rates (SDRs) for undetermined intent-to-suicide ratio and for undetermined intent-to-unintentional deaths in Portugal by sex, 2002–2023.

The sex-disaggregated SDR ratios confirm an important asymmetry throughout: the UnD:Suic ratio was consistently higher in females than in males (female peak: 2.25 in 2009 versus male peak: 1.08 in 2006), indicating that the relative weight of undetermined deaths on suicide certification was substantially greater in women.

The age-specific ratio analyses (Table 2, supplementary Tables S9–S10) reveal further heterogeneity. For the UnD:Suic ratio, the overall AAPC was negative or near zero in most male age groups. However, among females aged 45–64, the AAPC for the UnD:Suic ratio was significantly positive (+7.73%; 95% CI: 5.03–11.48), meaning undetermined deaths grew relative to suicide in this subgroup throughout the period, a sustained and widening misclassification dynamic specifically concentrated in middle-aged women.

The UnD:Accs ratio followed a broadly convergent pattern but with notable differences by age group. Among males aged 15–24, the AAPC for the UnD:Accs ratio was significantly positive (+4.66%; 95% CI: 1.92–8.27), absent from the UnD:Suic series for this group (AAPC: −1.58, non-significant), suggesting that in young males, undetermined deaths grew relative to accidents rather than relative to suicide, a different misclassification pathway. Among females aged 25–44 and 45–64, both ratios showed significantly positive AAPCs for the UnD:Accs series (+5.23% and +10.77% respectively), with the 45–64 group showing the most pronounced increase of any subgroup in any ratio (AAPC: +10.77%; 95% CI: 7.83– 13.74). By contrast, both ratios declined significantly after 2019 in most male age groups and in females aged 65+, consistent with the pandemic-era improvement in classification accuracy (see Fig. S2 of the supplementary file).

## 4. Discussion

The overall trends observed in this study show a decline in age-standardised death rates (SDRs) for suicide, undetermined intent deaths, and all unintentional deaths between 2002 and 2023 in Portugal, for both sexes. These patterns reflect improvements in mortality registration and waning misclassification over time, as well as probable actual changes in external cause mortality. The novel ratio analyses extend this picture by quantifying the dynamic relation between undetermined deaths and both suicide and accidents, disaggregated by sex and age group, revealing a more complex and subgroup-specific pattern of misclassification than aggregate trends alone would suggest.

Nevertheless, stakeholders and researchers continue to endorse the assumption that longitudinal suicide variation is constant - perpetuating and amplifying errors in the perception of the problem and in public policies related to suicide mitigation [33-36].

After the suspension in 2004 of the death certificate verification procedure implemented in 2002–2003 by the Directorate General of Health [2], suicide and unintentional deaths declined in both sexes, while undetermined deaths rose significantly, a pattern clearly visible in both the SDR trends and the ratio series, which peaked in 2005–2009. After 2005, undetermined deaths began to decline, while a period of stabilisation or slight increase in suicide from 2005 to 2014 could be partly attributed to the global financial crisis [17,18]. However, in agreement with Clement et al. [37], an alternative explanatory hypothesis is that heightened media coverage of suicide during the austerity period may have sensitised death certifiers, reducing stigma-driven undercounting and improving registry accuracy.

According to the literature, the global financial crisis and austerity from 2010 to 2014 would not have predicted an increase in all unintentional deaths [38,39], suggesting that the digitalisation of death certification (SICO) between 2013 and 2015 [15,16] further shaped trends, potentially improving classification in suicide and undetermined intent but not affecting unintentional deaths. This may have made it structurally harder to register uncertain causes as ‘undetermined’, thereby redirecting misclassified suicides into accidental categories rather than eliminating misclassification altogether. The decline in both ratio series after 2013–2015 is consistent with this interpretation - but the simultaneous rise in accident SDRs suggests the redirection was at least partly toward accidental rather than suicide categories, a dynamic only revealed by tracking both ratios simultaneously.

During the COVID-19 pandemic (2020–2021), undetermined deaths dropped sharply and both ratio indices reached their lowest values across the entire study period, consistent with the enhanced scrutiny of all deaths required for excess mortality reporting [19]. Registered suicide also reached its lowest levels. Many countries reported initial suicide declines during COVID-19 [40,41], and Portugal followed this pattern, though suicide rebounded in 2022 and the 2023 data suggest this rebound partially persisted. The partial recovery of undetermined deaths and both ratio indices in 2023 suggests that pandemic-era classification improvement was not sustained beyond the period of heightened scrutiny.

Age- and sex-specific trends reveal that suicide rates declined most in males aged 25–44, followed by 65+ and 45–64. The decline was statistically significant among females only in the oldest age group. Trends in undetermined deaths were less consistent, but significant reductions were observed in older age groups. Unintentional deaths decreased in all age groups except 65+, where an increase was significant only in females.

The ratio analyses identify a more precise picture of where misclassification is concentrated. The sustained and significantly positive AAPC of the UnD:Suic ratio among females aged 45–64 (+7.73%) signals a growing disproportion between undetermined deaths and suicide in this subgroup, running counter to the improving trend observed elsewhere. This is compounded by the even more pronounced positive AAPC for the UnD:Accs ratio in the same group (+10.77%), indicating that undetermined deaths are increasing relative to both reference categories simultaneously in middle-aged women. Among males aged 15–24, a significant positive AAPC for the UnD:Accs ratio (+4.66%) in the absence of a corresponding signal in the UnD:Suic series suggests a different misclassification pathway, possibly reflecting greater ambiguity in accident versus undetermined classification in younger males.

Identified joinpoints and breakpoints correspond to systemic events affecting data quality: the verbal autopsy period (2002–2004), data reporting disruptions (2004–2006), austerity (2009– 2014), digitalisation (2013–2015), and the pandemic (2020–2022). The coherence between breakpoints identified in the main SDR series and those in the ratio series reinforces this interpretation.

As in other countries, males consistently showed higher suicide rates across all age groups. The higher UnD:Suic ratios in females throughout the period are consistent with sex differences in method lethality and certifier behaviour: uncertain cases are more likely to be redirected away from suicide in women [5,6,13]. Underreporting affects male suicide cases in absolute terms, but in relative terms, the misclassification signal is stronger in females, particularly in middle age.

Taken together, the ratio analyses indicate that undetermined deaths in Portugal function simultaneously as a reservoir for concealed suicides and for misclassified accidents, and that this dual function varies substantially by sex and age group. The concentration of increasing misclassification signals in middle-aged women, and in young males for the UnD:Accs ratio, identifies these as the subgroups where surveillance improvements and forensic review protocols would have the greatest impact. These findings also provide the age- and sex-specific empirical foundation needed for future imputation-based estimation of probable suicide burden.

## 5. Strengths and limitations

This study provides a comprehensive national analysis of suicide and related mortality in Portugal over 22 years, integrating multiple external cause categories with robust trend and structural analysis methods. The novel dual-ratio approach, disaggregated by sex and four age groups, operationalises the misclassification hypothesis quantitatively.

We did not present the sum of suicide and undetermined death SDRs and ASDRs because, as the ratio data demonstrate, undetermined deaths in Portugal function as a reservoir for both misclassified suicides and misclassified accidents; simple summation would therefore exceedingly overestimate probable suicide burden. Future research should apply imputation-based methods using the ratio series as anchors, an approach that the current age- and sex-specific ratio data are specifically designed to support.

Limitations include reliance on secondary mortality data, which are subject to misclassification, coding inconsistencies, and regional variation in certification practices. We could not directly verify misclassified cases or establish causal attribution for individual breakpoints.

## 6. Conclusions

Registered suicide, undetermined intent, and unintentional deaths declined in Portugal from 2002 to 2023. However, divergent trends across age and sex subgroups, and their systematic correspondence with documented registry events, are consistent with ongoing suicide concealment and variable classification quality over time. The ratio analyses identify middle-aged women as the subgroup with the most pronounced and increasing misclassification signal, and post-2019 older age groups and males as showing the greatest recent improvement in classification accuracy.

These findings reinforce the need for (a) improved mortality registration systems and systematic forensic review protocols; (b) age- and sex-sensitive suicide prevention strategies, particularly for males and older adults; and (c) multi-sectoral surveillance approaches that account for systemic and demographic vulnerabilities in death classification.

Whenever registered suicides declined, increases in undetermined or accidental deaths were often observed, especially during periods of registry instability, supporting the hypothesis of masked suicide; homicides and traffic deaths declined during the same period, making misclassification into those categories unlikely [23].

## Data Availability

All data produced in the present study are available upon reasonable request to the authors.

## Acknowledgements

The authors have no acknowledgements to report.

