## Supplementary file for "Suicide trends in Portugal from 2002–2023 - a time-series analysis pondering data structure and fluctuations of undetermined intent and accidental deaths"

| **Tables & Figures** | **page** |
| --- | --- |
| Sup. Table S1: Number of deaths by suicide (X60-84) and SDR by sex, with male-to-female ratio, in Portugal, 2002-2023 (INE) | **2** |
| Sup. Table S2: Number of deaths by undetermined intent (Y10-34) and SDR by sex, with male-to-female ratio, in Portugal, 2002-2023 (INE) | **3** |
| Sup. Table S3: Number of deaths by accidents (V01-X59) and SDR by sex, with male-to-female ratio, in Portugal, 2002-2023 (INE) | **4** |
| Sup. Table S4: Undetermined-to-suicide SDR ratios (UnD:Suic), by sex, in Portugal, between 2002-2023 | **5** |
| Sup. Table S5: Undetermined-to-Accidents SDR ratios (UnD:Accs), overall and by sex, in Portugal, between 2002-2023 | **6** |
| Sup. Table S6: Age-specific death rates (ASDR, 15-24, 25-64, 65+ years) for suicide (X60-X84) by sex, in Portugal, 2002-2023 | **7** |
| Sup. Table S7: Age-specific death rates (ASDR, 15-24, 25-64, 65+ years) for undetermined intent deaths (Y10-34), total and by sex, in Portugal, 2002-2023 | **8** |
| Sup. Table S8: Age-specific death rates (ASDR, 15-24, 25-64, 65+ years) for unintentional deaths (V01-X59), by sex, in Portugal, 2002-2023 | **9** |
| Sup. Table S9: Undetermined-to-suicide ratios (UnD:Suic) for age-specific death rates (ASDR, 15-24, 25-64, 65+ years) by sex, in Portugal, 2002-2023 | **10** |
| Sup. Table S10: Undetermined-to-accident ratios (UnD:Accs) for age-specific death rates (ASDR, 15-24, 25-64, 65+ years) by sex, in Portugal, 2002-2023 | **11** |
| Sup. Table S11: Joinpoint regression for suicide SDR, undetermined intent SDR, and unintentional deaths SDR by sex in Portugal, 2002–2023 | **12** |
| Sup. Table S12: OLS segment regression results for structural breakpoints for suicide, undetermined deaths, and unintentional deaths by sex, Portugal, 2002–2023 | **13** |
| Sup. Table S13: Summary of segmented time trends by Joinpoint (JP) and Breakpoint (Bk) analyses for suicide, undetermined intent, and unintentional deaths by sex in Portugal, 2002–2023 (INE) | **14** |
| Figure S1: Trends in age-standardised death rates (SDRs) for suicide, undetermined intent, and unintentional deaths in Portugal by sex, 2002–2023 | **15** |
| Figure S2: Structural breakpoints in age-standardised death rates (SDRs) for undetermined intent-to-suicide ratio and for undetermined intent-to-unintentional deaths in Portugal by sex, 2002–2023 | **16** |

**Sup. Table S1: Number of deaths by suicide (X60-84) and SDR by sex, with male-to-female ratio, in Portugal, 2002-2023 (INE)**

|  | ****Number Suicide**** | | | ****SDR Suicide**** | | | |
| --- | --- | --- | --- | --- | --- | --- | --- |
| ****Year**** | ****Total**** | ****Male**** | ****Female**** | ****Total**** | ****Male**** | ****Female**** | ****Ratio M:F**** |
| 2002 | 1199 | 935 | 264 | 12.43 | 22.01 | 5.05 | 4,4 |
| 2003 | 1147 | 882 | 265 | 11.83 | 20.75 | 4.99 | 4,2 |
| 2004 | 1195 | 901 | 294 | 12.13 | 20.50 | 5.51 | 3,7 |
| 2005 | 910 | 692 | 218 | 9.16 | 15.68 | 4.05 | 3,9 |
| 2006 | 868 | 674 | 194 | 8.66 | 15.22 | 3.58 | 4,3 |
| 2007 | 1014 | 747 | 267 | 10.03 | 16.91 | 4.84 | 3,5 |
| 2008 | 1035 | 791 | 244 | 10.13 | 17.62 | 4.37 | 4,0 |
| 2009 | 1014 | 793 | 221 | 9.82 | 17.48 | 3.89 | 4,5 |
| 2010 | 1098 | 833 | 265 | 10.46 | 18.12 | 4.63 | 3,9 |
| 2011 | 1012 | 788 | 224 | 9.51 | 16.75 | 3.87 | 4,3 |
| 2012 | 1066 | 851 | 215 | 9.94 | 17.93 | 3.65 | 4,9 |
| 2013 | 1051 | 810 | 241 | 9.74 | 16.82 | 4.11 | 4,1 |
| 2014 | 1216 | 919 | 297 | 11.18 | 18.94 | 5.06 | 3,7 |
| 2015 | 1127 | 855 | 272 | 10.28 | 17.67 | 4.49 | 3,9 |
| 2016 | 975 | 746 | 229 | 8.88 | 15.20 | 3.82 | 4,0 |
| 2017 | 1048 | 778 | 270 | 9.51 | 15.73 | 4.55 | 3,5 |
| 2018 | 989 | 733 | 256 | 8.92 | 14.75 | 4.22 | 3,5 |
| 2019 | 975 | 738 | 237 | 8.64 | 14.58 | 3.87 | 3,8 |
| 2020 | 941 | 733 | 208 | 8.32 | 14.39 | 3.34 | 4,3 |
| 2021 | 928 | 720 | 208 | 8.12 | 13.92 | 3.41 | 4,1 |
| 2022 | 985 | 744 | 241 | 8.59 | 14.16 | 3.98 | 3,6 |
| 2023 | 1035 | 828 | 207 | 8.87 | 15.49 | 3.35 | 4.6 |
| **total** | 22828 | 17491 | 5337 |  |  |  |  |
| **variation %** |  |  |  | -28,64 | -29,62 | -33,66 |  |
| **average** |  |  |  | 9.78 | 16.85 | 4.21 | 4.0 |

**Sup. Table S2: Number of deaths by undetermined intent (Y10-34) and SDR by sex, with male-to-female ratio, in Portugal, 2002-2023 (INE)**

|  | **Number Undetermined** | | | | **SDR Undetermined** | | |  |
| --- | --- | --- | --- | --- | --- | --- | --- | --- |
| **Year** | | **Total** | **Men** | **Women** | **Total** | **Men** | **Women** | ****Ratio M:F**** |
| 2002 | | 288 | 214 | 74 | 3,00 | 4,88 | 1,42 | 3,4 |
| 2003 | | 322 | 226 | 96 | 3,37 | 5,24 | 1,85 | 2,8 |
| 2004 | | 771 | 518 | 253 | 8,13 | 12,06 | 4,82 | 2,5 |
| 2005 | | 988 | 665 | 323 | 10,48 | 15,88 | 6,12 | 2,6 |
| 2006 | | 1051 | 695 | 356 | 11,04 | 16,41 | 6,57 | 2,5 |
| 2007 | | 1054 | 620 | 434 | 11,22 | 15,29 | 7,88 | 1,9 |
| 2008 | | 1135 | 664 | 471 | 11,87 | 16,34 | 8,28 | 2,0 |
| 2009 | | 1145 | 635 | 510 | 11,70 | 15,33 | 8,75 | 1,8 |
| 2010 | | 1106 | 628 | 478 | 11,10 | 15,12 | 7,95 | 1,9 |
| 2011 | | 963 | 531 | 432 | 9,28 | 12,28 | 6,75 | 1,8 |
| 2012 | | 1007 | 586 | 421 | 9,35 | 13,22 | 6,35 | 2,1 |
| 2013 | | 865 | 480 | 385 | 7,90 | 10,67 | 5,67 | 1,9 |
| 2014 | | 878 | 490 | 388 | 7,88 | 10,65 | 5,66 | 1,9 |
| 2015 | | 778 | 465 | 313 | 6,92 | 9,92 | 4,51 | 2,2 |
| 2016 | | 654 | 430 | 224 | 5,79 | 8,87 | 3,23 | 2,7 |
| 2017 | | 639 | 412 | 227 | 5,57 | 8,43 | 3,25 | 2,6 |
| 2018 | | 799 | 474 | 325 | 6,79 | 9,67 | 4,46 | 2,2 |
| 2019 | | 1195 | 624 | 571 | 9,62 | 12,64 | 7,13 | 1,8 |
| 2020 | | 790 | 431 | 359 | 6,28 | 8,54 | 4,46 | 1,9 |
| 2021 | | 255 | 170 | 85 | 2,17 | 3,27 | 1,24 | 2,6 |
| 2022 | | 201 | 139 | 62 | 1,78 | 2,70 | 0,92 | 2,9 |
| 2023 | | 269 | 173 | 96 | 2,29 | 3,27 | 1,42 | 2,3 |
| **total** | | 17155 | 10270 | 6883 |  |  |  |  |
| **variation %** | |  |  |  | -23,67 | -32,99 | 0,00 |  |
| **average** | |  |  |  | 7,43 | 10,49 | 4,94 | 2,29 |

**Sup. Table S3: Number of deaths by accidents (V01-X59) and SDR by sex, with male-to-female ratio, in Portugal, 2002-2023 (INE)**

| **Number Unintentional** | | | | **SDR Unintentional** | | | |
| --- | --- | --- | --- | --- | --- | --- | --- |
| **Year** | **Total** | **Men** | **Women** | **Total** | **Men** | **Women** | ****Ratio M:F**** |
| 2002 | 3905 | 2827 | 1078 | 40,19 | 61,90 | 21,05 | 2,9 |
| 2003 | 3846 | 2695 | 1151 | 39,93 | 60,27 | 22,22 | 2,7 |
| 2004 | 3178 | 2285 | 893 | 32,19 | 49,43 | 17,03 | 2,9 |
| 2005 | 2372 | 1742 | 630 | 23,80 | 37,48 | 11,76 | 3,2 |
| 2006 | 2344 | 1699 | 645 | 23,44 | 37,20 | 11,84 | 3,1 |
| 2007 | 2041 | 1504 | 537 | 20,20 | 32,37 | 9,77 | 3,3 |
| 2008 | 1957 | 1413 | 544 | 19,22 | 30,43 | 9,71 | 3,1 |
| 2009 | 1952 | 1427 | 525 | 18,96 | 30,62 | 9,19 | 3,3 |
| 2010 | 1953 | 1358 | 595 | 18,78 | 28,88 | 10,15 | 2,8 |
| 2011 | 1788 | 1269 | 519 | 16,93 | 26,68 | 8,59 | 3,1 |
| 2012 | 1526 | 1056 | 470 | 14,26 | 22,34 | 7,50 | 3,0 |
| 2013 | 1996 | 1345 | 651 | 18,47 | 28,90 | 10,05 | 2,9 |
| 2014 | 2311 | 1518 | 793 | 21,10 | 32,45 | 11,86 | 2,7 |
| 2015 | 2530 | 1623 | 907 | 22,72 | 34,50 | 13,09 | 2,6 |
| 2016 | 2798 | 1705 | 1093 | 24,64 | 36,14 | 15,37 | 2,4 |
| 2017 | 3195 | 1924 | 1271 | 27,64 | 40,21 | 17,61 | 2,3 |
| 2018 | 3069 | 1865 | 1204 | 26,03 | 38,35 | 16,01 | 2,4 |
| 2019 | 2492 | 1525 | 967 | 21,16 | 30,71 | 13,16 | 2,3 |
| 2020 | 2985 | 1789 | 1196 | 24,44 | 35,43 | 15,32 | 2,3 |
| 2021 | 3695 | 2193 | 1502 | 29,50 | 42,66 | 18,74 | 2,3 |
| 2022 | 3869 | 2304 | 1565 | 30,57 | 44,05 | 19,42 | 2,3 |
| 2023 | 3892 | 2356 | 1536 | 30,13 | 44,07 | 18,68 | 2,4 |
| **total** | 59694 | 39422 | 20272 |  |  |  |  |
| **variation %** |  |  |  | -25,03 | -28,80 | -11,24 |  |
| **average** |  |  |  | 24,74 | 37,50 | 14,01 | 2,75 |

**Sup. Table S4: Undetermined-to-suicide SDR ratios (UnD:Suic), by sex, in Portugal, between 2002-2023**

|  | ****SDR Suic**** | | | ****SDR UnD**** | | | ****Ratio Und:Suic**** | | |
| --- | --- | --- | --- | --- | --- | --- | --- | --- | --- |
| ****Year**** | ****Total**** | ****Male**** | ****Female**** | ****Total**** | ****Male**** | ****Female**** | ****Total**** | ****Male**** | ****Female**** |
| 2002 | 12.43 | 22.01 | 5.05 | 3,00 | 4,88 | 1,42 | 0,24 | 0,22 | 0,28 |
| 2003 | 11.83 | 20.75 | 4.99 | 3,37 | 5,24 | 1,85 | 0,29 | 0,25 | 0,37 |
| 2004 | 12.13 | 20.50 | 5.51 | 8,13 | 12,06 | 4,82 | 0,67 | 0,59 | 0,88 |
| 2005 | 9.16 | 15.68 | 4.05 | 10,48 | 15,88 | 6,12 | 1,14 | 1,01 | 1,51 |
| 2006 | 8.66 | 15.22 | 3.58 | 11,04 | 16,41 | 6,57 | 1,28 | 1,08 | 1,83 |
| 2007 | 10.03 | 16.91 | 4.84 | 11,22 | 15,29 | 7,88 | 1,12 | 0,90 | 1,63 |
| 2008 | 10.13 | 17.62 | 4.37 | 11,87 | 16,34 | 8,28 | 1,17 | 0,93 | 1,89 |
| 2009 | 9.82 | 17.48 | 3.89 | 11,70 | 15,33 | 8,75 | 1,19 | 0,88 | 2,25 |
| 2010 | 10.46 | 18.12 | 4.63 | 11,10 | 15,12 | 7,95 | 1,06 | 0,83 | 1,72 |
| 2011 | 9.51 | 16.75 | 3.87 | 9,28 | 12,28 | 6,75 | 0,98 | 0,73 | 1,74 |
| 2012 | 9.94 | 17.93 | 3.65 | 9,35 | 13,22 | 6,35 | 0,94 | 0,74 | 1,74 |
| 2013 | 9.74 | 16.82 | 4.11 | 7,90 | 10,67 | 5,67 | 0,81 | 0,63 | 1,38 |
| 2014 | 11.18 | 18.94 | 5.06 | 7,88 | 10,65 | 5,66 | 0,70 | 0,56 | 1,12 |
| 2015 | 10.28 | 17.67 | 4.49 | 6,92 | 9,92 | 4,51 | 0,67 | 0,56 | 1,01 |
| 2016 | 8.88 | 15.20 | 3.82 | 5,79 | 8,87 | 3,23 | 0,65 | 0,58 | 0,85 |
| 2017 | 9.51 | 15.73 | 4.55 | 5,57 | 8,43 | 3,25 | 0,59 | 0,54 | 0,71 |
| 2018 | 8.92 | 14.75 | 4.22 | 6,79 | 9,67 | 4,46 | 0,76 | 0,66 | 1,05 |
| 2019 | 8.64 | 14.58 | 3.87 | 9,62 | 12,64 | 7,13 | 1,11 | 0,87 | 1,84 |
| 2020 | 8.32 | 14.39 | 3.34 | 6,28 | 8,54 | 4,46 | 0,75 | 0,59 | 1,33 |
| 2021 | 8.12 | 13.92 | 3.41 | 2,17 | 3,27 | 1,24 | 0,27 | 0,23 | 0,36 |
| 2022 | 8.59 | 14.16 | 3.98 | 1,78 | 2,70 | 0,92 | 0,21 | 0,19 | 0,23 |
| 2023 | 8.87 | 15.49 | 3.35 | 2,29 | 3,27 | 1,42 | 0,26 | 0,21 | 0,42 |

**Sup. Table S5: Undetermined-to-Accidents SDR ratios (UnD:Accs), overall and by sex, in Portugal, between 2002-2023**

|  | ****SDR Accs**** | | | ****SDR UnD**** | | | ****Ratio UnD:Accs**** | | |
| --- | --- | --- | --- | --- | --- | --- | --- | --- | --- |
| ****Year**** | ****Total**** | ****Male**** | ****Female**** | ****Total**** | ****Male**** | ****Female**** | ****Total**** | ****Male**** | ****Female**** |
| 2002 | 40,19 | 61,90 | 21,05 | 3,00 | 4,88 | 1,42 | 0,07 | 0,08 | 0,07 |
| 2003 | 39,93 | 60,27 | 22,22 | 3,37 | 5,24 | 1,85 | 0,08 | 0,09 | 0,08 |
| 2004 | 32,19 | 49,43 | 17,03 | 8,13 | 12,06 | 4,82 | 0,25 | 0,24 | 0,28 |
| 2005 | 23,80 | 37,48 | 11,76 | 10,48 | 15,88 | 6,12 | 0,44 | 0,42 | 0,52 |
| 2006 | 23,44 | 37,20 | 11,84 | 11,04 | 16,41 | 6,57 | 0,47 | 0,44 | 0,56 |
| 2007 | 20,20 | 32,37 | 9,77 | 11,22 | 15,29 | 7,88 | 0,56 | 0,47 | 0,81 |
| 2008 | 19,22 | 30,43 | 9,71 | 11,87 | 16,34 | 8,28 | 0,62 | 0,54 | 0,85 |
| 2009 | 18,96 | 30,62 | 9,19 | 11,70 | 15,33 | 8,75 | 0,62 | 0,50 | 0,95 |
| 2010 | 18,78 | 28,88 | 10,15 | 11,10 | 15,12 | 7,95 | 0,59 | 0,52 | 0,78 |
| 2011 | 16,93 | 26,68 | 8,59 | 9,28 | 12,28 | 6,75 | 0,55 | 0,46 | 0,79 |
| 2012 | 14,26 | 22,34 | 7,50 | 9,35 | 13,22 | 6,35 | 0,66 | 0,59 | 0,85 |
| 2013 | 18,47 | 28,90 | 10,05 | 7,90 | 10,67 | 5,67 | 0,43 | 0,37 | 0,56 |
| 2014 | 21,10 | 32,45 | 11,86 | 7,88 | 10,65 | 5,66 | 0,37 | 0,33 | 0,48 |
| 2015 | 22,72 | 34,50 | 13,09 | 6,92 | 9,92 | 4,51 | 0,30 | 0,29 | 0,34 |
| 2016 | 24,64 | 36,14 | 15,37 | 5,79 | 8,87 | 3,23 | 0,23 | 0,25 | 0,21 |
| 2017 | 27,64 | 40,21 | 17,61 | 5,57 | 8,43 | 3,25 | 0,20 | 0,21 | 0,18 |
| 2018 | 26,03 | 38,35 | 16,01 | 6,79 | 9,67 | 4,46 | 0,26 | 0,25 | 0,28 |
| 2019 | 21,16 | 30,71 | 13,16 | 9,62 | 12,64 | 7,13 | 0,45 | 0,41 | 0,54 |
| 2020 | 24,44 | 35,43 | 15,32 | 6,28 | 8,54 | 4,46 | 0,26 | 0,24 | 0,29 |
| 2021 | 29,50 | 42,66 | 18,74 | 2,17 | 3,27 | 1,24 | 0,07 | 0,08 | 0,07 |
| 2022 | 30,57 | 44,05 | 19,42 | 1,78 | 2,70 | 0,92 | 0,06 | 0,06 | 0,05 |
| 2023 | 30,13 | 44,07 | 18,68 | 2,29 | 3,27 | 1,42 | 0,08 | 0,07 | 0,08 |

**Sup. Table S6: Age-specific death rates (ASDR, 15-24, 25-44, 45-64, 65+ years) for suicide (X60-X84) by sex, in Portugal, 2002-2023**

| **Year** | **Total** | | | | **Men** | | | | **Women** | | | |
| --- | --- | --- | --- | --- | --- | --- | --- | --- | --- | --- | --- | --- |
|  | **15-24** | **25-44** | **45-64** | **65** | **15-24** | **25-44** | **45-64** | **65** | **15-24** | **25-44** | **45-64** | **65** |
| 2002 | 4,25 | 9,82 | 14,79 | 26,44 | 6,98 | 16,26 | 22,87 | 49,80 | 1,44 | 3,50 | 7,42 | 9,77 |
| 2003 | 3,59 | 9,23 | 14,40 | 25,37 | 5,62 | 14,67 | 22,49 | 47,19 | 1,49 | 3,91 | 7,02 | 9,82 |
| 2004 | 3,24 | 9,73 | 13,99 | 27,17 | 5,20 | 15,40 | 21,55 | 48,63 | 1,23 | 4,20 | 7,09 | 11,87 |
| 2005 | 2,95 | 7,19 | 10,26 | 20,89 | 5,35 | 11,61 | 15,71 | 37,25 | 0,47 | 2,88 | 5,28 | 9,25 |
| 2006 | 3,11 | 5,96 | 9,90 | 20,59 | 5,49 | 9,76 | 15,52 | 37,89 | 0,65 | 2,25 | 4,76 | 8,29 |
| 2007 | 2,94 | 6,39 | 12,10 | 24,37 | 4,17 | 9,73 | 18,32 | 43,68 | 1,66 | 3,13 | 6,41 | 10,67 |
| 2008 | 2,83 | 7,12 | 11,83 | 24,20 | 4,42 | 11,67 | 18,87 | 43,24 | 1,19 | 2,71 | 5,38 | 10,70 |
| 2009 | 4,08 | 6,50 | 12,56 | 21,78 | 6,69 | 10,60 | 20,79 | 39,80 | 1,38 | 2,54 | 5,02 | 9,01 |
| 2010 | 3,19 | 7,56 | 12,68 | 24,16 | 4,25 | 11,74 | 19,98 | 44,74 | 2,10 | 3,52 | 5,99 | 9,61 |
| 2011 | 2,10 | 7,15 | 13,37 | 19,50 | 2,59 | 11,15 | 21,80 | 37,42 | 1,60 | 3,31 | 5,65 | 6,83 |
| 2012 | 3,01 | 7,12 | 12,85 | 22,39 | 4,54 | 12,44 | 22,13 | 40,86 | 1,44 | 2,05 | 4,39 | 9,31 |
| 2013 | 2,24 | 7,10 | 13,01 | 21,71 | 3,53 | 11,89 | 20,96 | 39,29 | 0,91 | 2,56 | 5,77 | 9,25 |
| 2014 | 3,25 | 9,06 | 15,04 | 23,31 | 4,98 | 14,85 | 22,79 | 42,98 | 1,47 | 3,57 | 8,02 | 9,34 |
| 2015 | 3,17 | 7,48 | 13,12 | 23,45 | 5,34 | 11,62 | 21,69 | 41,41 | 0,92 | 3,57 | 5,38 | 10,63 |
| 2016 | 2,54 | 6,60 | 12,31 | 18,71 | 3,39 | 11,16 | 20,12 | 33,06 | 1,66 | 2,28 | 5,27 | 8,41 |
| 2017 | 3,83 | 7,33 | 13,40 | 18,80 | 5,91 | 11,45 | 20,22 | 33,91 | 1,68 | 3,42 | 7,26 | 7,88 |
| 2018 | 2,47 | 6,66 | 12,55 | 18,47 | 3,41 | 11,03 | 19,37 | 31,88 | 1,50 | 2,52 | 6,42 | 8,73 |
| 2019 | 3,39 | 6,15 | 12,36 | 17,66 | 5,57 | 10,08 | 19,20 | 31,38 | 1,12 | 2,42 | 6,23 | 7,60 |
| 2020 | 3,94 | 6,96 | 10,63 | 16,89 | 5,92 | 12,21 | 17,09 | 30,12 | 1,87 | 1,93 | 4,84 | 7,08 |
| 2021 | 3,21 | 6,11 | 11,53 | 16,09 | 4,66 | 9,82 | 18,47 | 29,61 | 1,69 | 2,51 | 5,30 | 5,96 |
| 2022 | 4,79 | 5,93 | 12,43 | 16,30 | 7,20 | 9,30 | 19,44 | 28,91 | 2,26 | 2,64 | 6,12 | 6,78 |
| 2023 | 4,29 | 7,11 | 12,60 | 16,55 | 6,62 | 11,50 | 21,04 | 31,00 | 1,86 | 2,78 | 5,00 | 5,61 |

**Sup. Table S7: Age-specific death rates (ASDR, 15-24, 25-44, 45-64, 65+ years) for undetermined intent deaths (Y10-34), total and by sex, in Portugal, 2002-2023**

| **Year** | **Total** | | | | **Men** | | | | **Women** | | | |
| --- | --- | --- | --- | --- | --- | --- | --- | --- | --- | --- | --- | --- |
|  | **15-24** | **25-44** | **45-64** | **65** | **15-24** | **25-44** | **45-64** | **65** | **15-24** | **25-44** | **45-64** | **65** |
| 2002 | 1,28 | 2,31 | 2,34 | 7,37 | 2,23 | 3,82 | 4,49 | 10,79 | 0,29 | 0,83 | 0,39 | 4,94 |
| 2003 | 1,83 | 2,26 | 3,11 | 7,89 | 3,03 | 4,05 | 4,85 | 11,05 | 0,59 | 0,50 | 1,53 | 5,64 |
| 2004 | 1,58 | 4,95 | 7,80 | 21,36 | 2,38 | 7,96 | 13,04 | 28,08 | 0,77 | 2,01 | 3,02 | 16,57 |
| 2005 | 3,58 | 5,00 | 7,81 | 30,59 | 5,66 | 8,79 | 12,61 | 41,49 | 1,42 | 1,31 | 3,42 | 22,84 |
| 2006 | 5,27 | 5,45 | 7,99 | 32,19 | 8,00 | 9,12 | 14,00 | 41,69 | 2,44 | 1,88 | 2,49 | 25,43 |
| 2007 | 2,45 | 3,81 | 7,21 | 37,71 | 3,69 | 6,25 | 12,19 | 44,07 | 1,17 | 1,44 | 2,66 | 33,19 |
| 2008 | 1,92 | 4,15 | 6,88 | 41,71 | 2,79 | 6,94 | 11,29 | 49,51 | 1,02 | 1,45 | 2,83 | 36,18 |
| 2009 | 2,46 | 3,35 | 7,68 | 41,37 | 3,85 | 5,56 | 11,81 | 46,10 | 1,04 | 1,20 | 3,91 | 38,02 |
| 2010 | 1,99 | 3,84 | 6,90 | 38,45 | 3,06 | 6,04 | 11,27 | 43,63 | 0,88 | 1,73 | 2,89 | 34,78 |
| 2011 | 1,31 | 2,42 | 6,01 | 34,94 | 2,24 | 4,19 | 10,86 | 37,05 | 0,35 | 0,71 | 1,57 | 33,45 |
| 2012 | 1,42 | 2,46 | 6,32 | 35,59 | 2,62 | 4,15 | 10,58 | 42,52 | 0,18 | 0,86 | 2,43 | 30,69 |
| 2013 | 0,81 | 2,14 | 5,34 | 30,80 | 1,06 | 3,54 | 8,56 | 35,32 | 0,55 | 0,81 | 2,42 | 27,59 |
| 2014 | 1,26 | 2,82 | 6,03 | 28,62 | 2,13 | 4,13 | 10,18 | 31,32 | 0,37 | 1,58 | 2,27 | 26,71 |
| 2015 | 1,36 | 3,45 | 6,24 | 22,61 | 1,96 | 5,40 | 10,22 | 26,97 | 0,74 | 1,61 | 2,66 | 19,49 |
| 2016 | 1,27 | 2,64 | 6,28 | 17,57 | 2,32 | 4,53 | 11,05 | 22,33 | 0,18 | 0,86 | 1,98 | 14,15 |
| 2017 | 1,37 | 2,65 | 5,61 | 17,09 | 1,79 | 4,69 | 9,31 | 22,25 | 0,93 | 0,73 | 2,29 | 13,36 |
| 2018 | 1,56 | 2,78 | 5,88 | 23,62 | 2,33 | 4,30 | 9,97 | 28,01 | 0,75 | 1,33 | 2,21 | 20,42 |
| 2019 | 1,10 | 2,90 | 6,00 | 39,83 | 1,44 | 5,00 | 9,89 | 41,74 | 0,75 | 0,91 | 2,51 | 38,43 |
| 2020 | 1,19 | 2,12 | 3,45 | 25,74 | 1,61 | 3,61 | 5,96 | 28,54 | 0,75 | 0,69 | 1,21 | 23,66 |
| 2021 | 0,55 | 1,12 | 2,99 | 5,29 | 1,08 | 1,62 | 4,49 | 7,57 | 0,01 | 0,63 | 1,64 | 3,59 |
| 2022 | 1,10 | 1,70 | 2,14 | 3,22 | 1,62 | 3,10 | 3,19 | 4,12 | 0,57 | 0,32 | 1,19 | 2,54 |
| 2023 | 0,91 | 1,52 | 3,15 | 4,73 | 1,43 | 2,33 | 4,73 | 5,76 | 0,37 | 0,71 | 1,73 | 3,95 |

**Sup. Table S8: Age-specific death rates (ASDR, 15-24, 25-44, 45-64, 65+ years) for unintentional deaths (V01-X59), by sex, in Portugal, 2002-2023**

| **Year** | **Total** | | | | **Male** | | | | **Female** | | | |
| --- | --- | --- | --- | --- | --- | --- | --- | --- | --- | --- | --- | --- |
|  | **15-24** | **25-44** | **45-64** | **65** | **15-24** | **25-44** | **45-64** | **65** | **15-24** | **25-44** | **45-64** | **65** |
| 2002 | 32,53 | 33,37 | 31,96 | 83,42 | 54,30 | 58,69 | 52,26 | 110,82 | 10,08 | 8,52 | 13,44 | 63,87 |
| 2003 | 30,51 | 28,17 | 32,27 | 91,22 | 50,45 | 48,38 | 55,09 | 117,84 | 9,96 | 8,38 | 11,45 | 72,25 |
| 2004 | 29,33 | 26,70 | 24,32 | 67,48 | 47,81 | 47,54 | 40,04 | 88,94 | 10,27 | 6,33 | 9,96 | 52,19 |
| 2005 | 23,63 | 18,94 | 19,79 | 49,34 | 39,29 | 32,71 | 34,10 | 67,47 | 7,43 | 5,51 | 6,70 | 36,43 |
| 2006 | 17,00 | 17,94 | 20,07 | 52,99 | 28,08 | 30,64 | 33,77 | 74,99 | 5,53 | 5,57 | 7,54 | 37,36 |
| 2007 | 17,80 | 15,66 | 16,76 | 44,32 | 29,34 | 27,96 | 28,23 | 62,99 | 5,83 | 3,70 | 6,26 | 31,08 |
| 2008 | 15,17 | 13,57 | 16,63 | 44,68 | 23,27 | 24,24 | 28,08 | 64,48 | 6,79 | 3,21 | 6,16 | 30,65 |
| 2009 | 14,19 | 13,94 | 16,96 | 43,30 | 22,24 | 24,28 | 29,10 | 63,11 | 5,87 | 3,93 | 5,86 | 29,27 |
| 2010 | 10,79 | 14,01 | 16,91 | 45,00 | 16,33 | 24,08 | 27,41 | 62,17 | 5,08 | 4,29 | 7,29 | 32,86 |
| 2011 | 12,50 | 11,22 | 16,82 | 40,19 | 18,96 | 20,07 | 27,53 | 56,73 | 5,86 | 2,73 | 7,02 | 28,50 |
| 2012 | 8,15 | 9,49 | 12,25 | 37,90 | 14,31 | 17,34 | 20,87 | 50,34 | 1,80 | 1,98 | 4,39 | 29,10 |
| 2013 | 9,41 | 9,62 | 15,43 | 55,11 | 15,18 | 17,26 | 25,17 | 76,01 | 3,46 | 2,36 | 6,58 | 40,31 |
| 2014 | 8,76 | 10,82 | 18,09 | 64,70 | 14,76 | 19,35 | 31,49 | 82,30 | 2,57 | 2,74 | 5,95 | 52,21 |
| 2015 | 7,69 | 11,14 | 17,37 | 74,40 | 13,17 | 20,13 | 30,58 | 93,34 | 2,02 | 2,66 | 5,44 | 60,88 |
| 2016 | 8,71 | 8,58 | 19,00 | 86,84 | 14,44 | 15,08 | 33,00 | 104,97 | 2,77 | 2,42 | 6,39 | 73,81 |
| 2017 | 8,67 | 12,11 | 20,15 | 96,76 | 12,71 | 21,05 | 34,70 | 115,54 | 4,47 | 3,64 | 7,06 | 83,20 |
| 2018 | 8,79 | 10,12 | 19,18 | 93,36 | 14,72 | 19,16 | 33,90 | 110,70 | 2,62 | 1,56 | 5,97 | 80,77 |
| 2019 | 8,99 | 10,14 | 16,90 | 69,27 | 12,93 | 18,17 | 28,37 | 82,77 | 4,87 | 2,49 | 6,62 | 59,37 |
| 2020 | 8,16 | 10,54 | 16,50 | 88,04 | 13,81 | 18,88 | 29,08 | 102,46 | 2,25 | 2,54 | 5,22 | 77,33 |
| 2021 | 8,71 | 10,14 | 20,93 | 110,30 | 14,53 | 17,37 | 37,08 | 129,46 | 2,63 | 3,14 | 6,43 | 95,95 |
| 2022 | 11,23 | 11,02 | 21,11 | 112,69 | 17,82 | 19,00 | 37,34 | 131,94 | 4,33 | 3,20 | 6,50 | 98,15 |
| 2023 | 9,77 | 11,78 | 20,43 | 111,79 | 16,65 | 21,08 | 34,96 | 135,14 | 2,61 | 2,62 | 7,34 | 94,11 |

**Sup. Table S9: Undetermined-to-suicide ratios (UnD:Suic) for age-specific death rates (ASDR, 15-24, 25-44, 45-64, 65+ years) by sex, in Portugal, 2002-2023**

| **Year** | **Total** | | | | **Men** | | | | **Women** | | | |
| --- | --- | --- | --- | --- | --- | --- | --- | --- | --- | --- | --- | --- |
|  | **15-24** | **25-44** | **45-64** | **65** | **15-24** | **25-44** | **45-64** | **65** | **15-24** | **25-44** | **45-64** | **65** |
| 2002 | 0,30 | 0,24 | 0,16 | 0,28 | 0,32 | 0,24 | 0,20 | 0,22 | 0,20 | 0,24 | 0,05 | 0,51 |
| 2003 | 0,51 | 0,24 | 0,22 | 0,31 | 0,54 | 0,28 | 0,22 | 0,23 | 0,40 | 0,13 | 0,22 | 0,57 |
| 2004 | 0,49 | 0,51 | 0,56 | 0,79 | 0,46 | 0,52 | 0,61 | 0,58 | 0,63 | 0,48 | 0,43 | 1,40 |
| 2005 | 1,21 | 0,70 | 0,76 | 1,46 | 1,06 | 0,76 | 0,80 | 1,11 | 3,00 | 0,46 | 0,65 | 2,47 |
| 2006 | 1,69 | 0,91 | 0,81 | 1,56 | 1,46 | 0,93 | 0,90 | 1,10 | 3,75 | 0,83 | 0,52 | 3,07 |
| 2007 | 0,83 | 0,60 | 0,60 | 1,55 | 0,88 | 0,64 | 0,67 | 1,01 | 0,70 | 0,46 | 0,42 | 3,11 |
| 2008 | 0,68 | 0,58 | 0,58 | 1,72 | 0,63 | 0,59 | 0,60 | 1,14 | 0,86 | 0,53 | 0,53 | 3,38 |
| 2009 | 0,60 | 0,51 | 0,61 | 1,90 | 0,58 | 0,52 | 0,57 | 1,16 | 0,75 | 0,48 | 0,78 | 4,22 |
| 2010 | 0,62 | 0,51 | 0,54 | 1,59 | 0,72 | 0,51 | 0,56 | 0,98 | 0,42 | 0,49 | 0,48 | 3,62 |
| 2011 | 0,63 | 0,34 | 0,45 | 1,79 | 0,87 | 0,38 | 0,50 | 0,99 | 0,22 | 0,22 | 0,28 | 4,90 |
| 2012 | 0,47 | 0,35 | 0,49 | 1,59 | 0,58 | 0,33 | 0,48 | 1,04 | 0,13 | 0,42 | 0,55 | 3,30 |
| 2013 | 0,36 | 0,30 | 0,41 | 1,42 | 0,30 | 0,30 | 0,41 | 0,90 | 0,60 | 0,32 | 0,42 | 2,98 |
| 2014 | 0,39 | 0,31 | 0,40 | 1,23 | 0,43 | 0,28 | 0,45 | 0,73 | 0,25 | 0,44 | 0,28 | 2,86 |
| 2015 | 0,43 | 0,46 | 0,48 | 0,96 | 0,37 | 0,46 | 0,47 | 0,65 | 0,80 | 0,45 | 0,49 | 1,83 |
| 2016 | 0,50 | 0,40 | 0,51 | 0,94 | 0,68 | 0,41 | 0,55 | 0,68 | 0,11 | 0,38 | 0,38 | 1,68 |
| 2017 | 0,36 | 0,36 | 0,42 | 0,91 | 0,30 | 0,41 | 0,46 | 0,66 | 0,56 | 0,21 | 0,32 | 1,70 |
| 2018 | 0,63 | 0,42 | 0,47 | 1,28 | 0,68 | 0,39 | 0,51 | 0,88 | 0,50 | 0,53 | 0,34 | 2,34 |
| 2019 | 0,32 | 0,47 | 0,48 | 2,26 | 0,26 | 0,50 | 0,51 | 1,33 | 0,67 | 0,38 | 0,40 | 5,06 |
| 2020 | 0,30 | 0,31 | 0,32 | 1,52 | 0,27 | 0,30 | 0,35 | 0,95 | 0,40 | 0,36 | 0,25 | 3,34 |
| 2021 | 0,17 | 0,18 | 0,26 | 0,33 | 0,23 | 0,17 | 0,24 | 0,26 | 0,01 | 0,25 | 0,31 | 0,60 |
| 2022 | 0,23 | 0,29 | 0,17 | 0,20 | 0,23 | 0,33 | 0,16 | 0,14 | 0,25 | 0,12 | 0,19 | 0,38 |
| 2023 | 0,21 | 0,21 | 0,25 | 0,29 | 0,22 | 0,20 | 0,22 | 0,19 | 0,20 | 0,26 | 0,35 | 0,70 |

**Sup. Table S10: Undetermined-to-accident ratios (UnD:Accs) for age-specific death rates (ASDR, 15-24, 25-44, 45-64, 65+ years) by sex, in Portugal, 2002-2023**

| **Year** | **Total** | | | | **Men** | | | | **Women** | | | |
| --- | --- | --- | --- | --- | --- | --- | --- | --- | --- | --- | --- | --- |
|  | **15-24** | **25-44** | **45-64** | **65** | **15-24** | **25-44** | **45-64** | **65** | **15-24** | **25-44** | **45-64** | **65** |
| 2002 | 0,04 | 0,07 | 0,11 | 0,09 | 0,04 | 0,07 | 0,09 | 0,10 | 0,03 | 0,10 | 0,03 | 0,08 |
| 2003 | 0,06 | 0,08 | 0,07 | 0,09 | 0,06 | 0,08 | 0,09 | 0,09 | 0,06 | 0,06 | 0,13 | 0,08 |
| 2004 | 0,05 | 0,19 | 0,10 | 0,32 | 0,05 | 0,17 | 0,33 | 0,32 | 0,07 | 0,32 | 0,30 | 0,32 |
| 2005 | 0,15 | 0,26 | 0,32 | 0,62 | 0,14 | 0,27 | 0,37 | 0,61 | 0,19 | 0,24 | 0,51 | 0,63 |
| 2006 | 0,31 | 0,30 | 0,39 | 0,61 | 0,28 | 0,30 | 0,41 | 0,56 | 0,44 | 0,34 | 0,33 | 0,68 |
| 2007 | 0,14 | 0,24 | 0,40 | 0,85 | 0,13 | 0,22 | 0,43 | 0,70 | 0,20 | 0,39 | 0,43 | 1,07 |
| 2008 | 0,13 | 0,31 | 0,43 | 0,93 | 0,12 | 0,29 | 0,40 | 0,77 | 0,15 | 0,45 | 0,46 | 1,18 |
| 2009 | 0,17 | 0,24 | 0,41 | 0,96 | 0,17 | 0,23 | 0,41 | 0,73 | 0,18 | 0,31 | 0,67 | 1,30 |
| 2010 | 0,18 | 0,27 | 0,45 | 0,85 | 0,19 | 0,25 | 0,41 | 0,70 | 0,17 | 0,40 | 0,40 | 1,06 |
| 2011 | 0,10 | 0,22 | 0,41 | 0,87 | 0,12 | 0,21 | 0,39 | 0,65 | 0,06 | 0,26 | 0,22 | 1,17 |
| 2012 | 0,17 | 0,26 | 0,36 | 0,94 | 0,18 | 0,24 | 0,51 | 0,84 | 0,10 | 0,43 | 0,55 | 1,05 |
| 2013 | 0,09 | 0,22 | 0,52 | 0,56 | 0,07 | 0,20 | 0,34 | 0,46 | 0,16 | 0,34 | 0,37 | 0,68 |
| 2014 | 0,14 | 0,26 | 0,35 | 0,44 | 0,14 | 0,21 | 0,32 | 0,38 | 0,14 | 0,58 | 0,38 | 0,51 |
| 2015 | 0,18 | 0,31 | 0,33 | 0,30 | 0,15 | 0,27 | 0,33 | 0,29 | 0,36 | 0,61 | 0,49 | 0,32 |
| 2016 | 0,15 | 0,31 | 0,36 | 0,20 | 0,16 | 0,30 | 0,33 | 0,21 | 0,07 | 0,35 | 0,31 | 0,19 |
| 2017 | 0,16 | 0,22 | 0,33 | 0,18 | 0,14 | 0,22 | 0,27 | 0,19 | 0,21 | 0,20 | 0,32 | 0,16 |
| 2018 | 0,18 | 0,27 | 0,28 | 0,25 | 0,16 | 0,22 | 0,29 | 0,25 | 0,29 | 0,86 | 0,37 | 0,25 |
| 2019 | 0,12 | 0,29 | 0,31 | 0,58 | 0,11 | 0,28 | 0,35 | 0,50 | 0,15 | 0,36 | 0,38 | 0,65 |
| 2020 | 0,15 | 0,20 | 0,35 | 0,29 | 0,12 | 0,19 | 0,20 | 0,28 | 0,33 | 0,27 | 0,23 | 0,31 |
| 2021 | 0,06 | 0,11 | 0,21 | 0,05 | 0,07 | 0,09 | 0,12 | 0,06 | 0,00 | 0,20 | 0,25 | 0,04 |
| 2022 | 0,10 | 0,15 | 0,14 | 0,03 | 0,09 | 0,16 | 0,09 | 0,03 | 0,13 | 0,10 | 0,18 | 0,03 |
| 2023 | 0,09 | 0,13 | 0,10 | 0,04 | 0,09 | 0,07 | 0,14 | 0,04 | 0,14 | 0,27 | 0,24 | 0,04 |

**Sup. Table S11: Joinpoint regression for suicide SDR, undetermined intent SDR, and unintentional deaths SDR by sex in Portugal, 2002–2023**

|  | **Male** | | | | | | **Female** | | | | |
| --- | --- | --- | --- | --- | --- | --- | --- | --- | --- | --- | --- |
|  |  | | **Period (JP)** | **Value**  **(95% CI)** | **B**  **(SE)** | **p-value** |  | **Period (JP)** | **Value**  **(95% CI)** | **B**  **(SE)** | **p-value** |
| **Suicide** | |  |  |  |  |  | **AAPC** | **None** | **-1.32**  **(-2.30; -0.28)** | **-** | **-** |
|  |  | **AAPC** | **(2006; 2009)** | **-2.25**  **(-3.02; -1.49)** | **-** | **-** |  |  |  |  |  |
|  |  | APC 1 | **2002-06** | -9.18  (-18.29; -3.63) | -0.10  (0.03) | <0.01 | APC 1 | **2002-23** | -1.32  (-2.30; -0.28) | -0.01  (0.00) | <0.01 |
|  |  | APC 2 | 2006-09 | 5.95  (-0.76; 10.94) | 0.06  (0.09) | 0.546 | - | - | - | - | - |
|  |  | APC 3 | **2009-23** | -1.88  (-4.81; -1.12) | -0.02  (0.00) | <0.01 | - | - | - | - | - |
| **Undetermined**  **deaths** | | **AAPC** | **(2005; 2020)** | **-2.61**  **(-5.01; -0.14)** | **-** | **-** | **AAPC** | **(2006; 2019)** | -2.05  (-5.54; 1.67) | **-** | **-** |
|  |  | APC 1 | **2002-05** | **59.01**  **(32.63; 115.75)** | 0.46  (0.11) | <0.001 | APC 1 | **2002-06** | 56.32  (27.17; 134.87) | 0.45 (0.12) | 0.003 |
|  |  | APC 2 | **2005-20** | **-4.65**  **(-6.44; -2.58)** | -0.05  (0.01) | <0.001 | APC 2 | 2006-19 | -5.59  (-9.68; 1.54) | -0.06 (0.02) | 0.025 |
|  |  | APC 3 | **2020-23** | **-47.38**  **(-57.49; -29.18)** | -0.64  (0.22) | <0.05 | APC 3 | **2019-23** | -30.81  (-54.57; -16.15) | -0.37 (0.12) | 0.010 |
| **Unintentional**  **deaths** | | **AAPC** | **(2010)** | **-1.38**  **(-2.27; -0.41)** | **-** | **-** | **AAPC** | (2008) | -0.66  (-2.03; 0.94) | **-** | **-** |
|  |  | APC 1 | **2002-10** | **-10.05**  **(-14.17; -7.09)** | -0.11  (0.02) | <0.001 | APC 1 | **2002-08** | -15.51  (-26.66; -9.04) | -0.17  (0.03) | <0.001 |
|  |  | APC 2 | **2010-23** | **4.37**  **(2.70; 6.52)** | 0.04  (0.01) | <0.001 | APC 2 | **2008-23** | 5.99  (4.03; 8.66) | 0.06  (0.01) | <0.001 |
| **Ratio undetermined-**  **to-suicide** | | **AAPC** | **(2005;**  **2019)** | **-0.50**  **(-3.30; 2.46)** | **-** | **-** | **AAPC** | **(2006)** | **3.16**  **(-0.47; 7.48)** | **-** | **-** |
|  |  | APC 1 | 2002-05 | 71.82  (37.53; 147.13) | 0.54  (0.13) | <0.01 | APC 1 | **2002-06** | 75.59  (32.48; 202.78) | 0.56  (0.18) | <0.01 |
|  |  | APC 2 | 2005-19 | -3.54  (-6.27; 0.03) | -0.04  (0.01) | <0.05 | APC 2 | **2006-23** | -8.97  (-12.84; -5.53) | -0.09  (0.02) | <0.05 |
|  |  | APC 3 | 2019-23 | -26.38  (-45.17; -15.42) | -0.31  (0.08) | <0.01 | - | - | - | - | - |
| **Ratio undetermined-**  **to-accidents** | | **AAPC** | **(2006; 2019)** | -1.35  (-5.26; 2.91) | **-** | **-** | **AAPC** | **(2008)** | 0.27  (-4.19; 6.08) | **-** | **-** |
|  |  | APC1 | 2002-06 | 68.96  (36.40; 158.61) | 0,52  (0,11) | <0.001 | APC1 | 2002-08 | 60.41  (28.78; 133.89) | 0,47  (0,11) | <0.001 |
|  |  | APC2 | 2006-19 | -6.04  (-9.97; 0.71) | -0,06 (0,02) | <0.05 | APC2 | 2008-23 | -16.91  (-22.60; -11.83) | -0,19  (0,03) | <0.001 |
|  |  | APC3 | 2019-23 | -32.52  (-56.91; -18.02) | -0,39 (0,11) | <0.01 |  |  |  |  |  |

Note: AAPC=average annual percent change; APC = annual percent change for each time segment; B = slope change coefficient; SE=standard error. Slope change coefficients reflect differences from the previous segment; the first segment is compared against a zero slope. Significant values are highlighted in bold; undt= undetermined deaths

**Sup. Table S12: OLS segment regression results for structural breakpoints for suicide, undetermined deaths, and unintentional deaths by sex, Portugal, 2002–2023**

| **Outcome** | **Sex** | **Segment (Years)** | **Breakpoint (95% CI)** | **Intercept** | **Slope** | **p-value (slope)** | **Combined Slope**  **at Start**  **(95% CI)** |
| --- | --- | --- | --- | --- | --- | --- | --- |
| **Suicide** | **Male** | 2002–2004 | – | 1526.44 | -0.75 | 0.149 | – |
|  |  | 2004–2007 | **2004**  (2003–2005) | -2738.59 | 1.36 | 0.070 | 0.61  (-0.35; 1.58) |
|  |  | 2007-2015 | **2007**  (2005–2009) | -1629.25 | 0.81 | 0.129 | 0.06  (-0.15; 0.27) |
|  |  | 2015-2023 | **2015**  (2014–2016) | -1306.22 | 0.65 | 0.217 | -0.10  (-0.31; 0.11) |
|  | **Female** | 2002–2004 | – | -453.55 | 0.23 | 0.462 | – |
|  |  | 2004–2018 | **2004**  (2001–2005) | 419.88 | -0.21 | 0.501 | 0.02  (-0.04; 0.07) |
|  |  | 2018–2023 | **2018**  (2016–2024) | 540.62 | -0.27 | 0.429 | -0.04  (-0.31; 0.23) |
| **Undetermined deaths** | **Male** | 2002–2004 | – | -7184.19 | 3.59 | <0.01 | – |
|  |  | 2004–2010 | **2004**  (2004–2004) | 7543.03 | -3.76 | <0.01 | -0.17  (-0.84; 0.49) |
|  |  | 2010–2020 | **2010**  (2008–2011) | 7811.06 | -3.90 | <0.01 | -0.31  (-0.61; 0.00) |
|  |  | 2020–2023 | **2020**  (2019–2021) | 7186.40 | -3.59 | <0.05 | 0.00  (-1.94; 1.95) |
|  | **Female** | 2002–2004 | – | -3397.64 | 1.70 | 0.065 | – |
|  |  | 2004–2012 | **2004**  (2003–2006) | 3329.38 | -1.66 | 0.077 | -0.17  (-0.89; 0.55) |
|  |  | 2012–2020 | **2012**  (2009–2014) | 3433.41 | -1.71 | 0.069 | -0.26  (-0.38; 0.35) |
|  |  | 2020–2023 | **2020**  (2019–2022) | 3220.04 | -1.61 | 0.201 | 0.09  (-1.57; 1.75) |
| **Unintentional**  **deaths** | **Male** | 2002–2004 | – | 12537.68 | -6.23 | <0.05 | – |
|  |  | 2004–2007 | **2004**  (2003–2005) | -7371.23 | 3.67 | 0.253 | -2.56  (-6.80; 1.68) |
|  |  | 2007–2013 | **2007**  (2005–2008) | -10518.84 | 5.24 | <0.05 | -0.99  (-2.42; 0.44) |
|  |  | 2013–2020 | **2013**  (2011–2015) | -12761.02 | 6.36 | <0.05 | 0.13  (-1.00; 1.26) |
|  |  | 2020–2023 | **2020**  (2019–2022) | -13917.93 | 6.94 | <0.05 | 0.70  (-3.53; 4.94) |
|  | **Female** | 2002–2004 | – | 4044.80 | -2.01 | 0.111 | – |
|  |  | 2004–2012 | **2004**  (2003–2005) | -3725.66 | 1.86 | 0.143 | -0.15  (-0.51; 0.21) |
|  |  | 2012–2019 | **2012**  (2011–2015) | -4198.84 | 2.09 | 0.115 | 0.08  (-0.70; 0.87) |
|  |  | 2019–2023 | **2019**  (2020–2023) | -3969.56 | 1.98 | 0.255 | -0.03  (-2.34; 2.29) |
| **Ratio**  **Undetermined-to-Suicide** | **Male** | 2002–2004 | – | -366.66 | 0.18 | <0.05 | – |
|  |  | 2004–2012 | **2004**  (2003–2005) | 451.90 | -0.23 | <0.01 | -0.04  (-0.09; 0.00) |
|  |  | 2012–2019 | **2010**  (2008–2011) | 371.01 | -0.19 | <0.05 | -0.00  (-0.02; 0.02) |
|  |  | 2019–2023 | 2020  (2019–2021) | 390.92 | -0.20 | 0.057 | -0.01  (-0.14; 0.12) |
|  | **Female** | 2002–2004 | – | -593.57 | 0.30 | 0.233 | – |
|  |  | 2004-2012 | 2004  (2003–2005) | 450.00 | -0.22 | 0.388 | 0.072  (-0.09; 0.23) |
|  |  | 2012-2020 | 2012  (2008–2013) | 667.51 | -0.33 | 0.189 | -0.04  (-0.11; 0.04) |
|  |  | 2020-2023 | 2020  (2019-2022 | 533.83 | -0.27 | 0.442 | 0.03  (-0.44; 0.50) |
| **Ratio**  **Undetermined-to-Accidents** | **Male** | 2002-2004 | - | -215.55 | 0.11 | 0.306 | - |
|  |  | 2004-2012 | 2004  (2003–2005) | 184.91 | -0.09 | 0.387 | 0.02  (-0.02; 0.05) |
|  |  | 2012-2020 | 2012  (2010–2013) | 222.82 | -0.11 | 0.308 | -0.00  (-0.06; 0.05) |
|  |  | 2020-2023 | 2020  (2020-2020) | 205.85 | -0.10 | 0.485 | 0.00  (-0.19; 0.20) |
|  | **Female** | 2002-2004 | - | -165.23 | 0.08 | 0.062 | - |
|  |  | 2004-2013 | 2004  (2003–2005) | 132.50 | -0.07 | 0.136 | 0.02  (-0.00; 0.03) |
|  |  | 2013-2020 | **2013**  (2012–2015) | 180.50 | -0.09 | <0.05 | -0.01  (-0.02; 0.01) |
|  |  | 2020-2023 | 2020  (2020-2020) | 167.76 | -0.08 | 0.168 | -0.00  (-0.08; 0.08) |

Note: Slope change coefficients reflect differences from the previous segment; the first segment is compared against a zero slope. CI= Confidence intervals. Significant values are highlighted in bold.

**Sup. Table S13: Summary of segmented time trends by Joinpoint (JP) and Breakpoint (Bk) analyses for suicide, undetermined intent, and unintentional deaths by sex in Portugal, 2002–2023 (INE)**

|  |  | **Suicide** |  | **UnD** |  | **Accidents** |  | **UnD:Suic** |  | **UnD:Accs** |  |
| --- | --- | --- | --- | --- | --- | --- | --- | --- | --- | --- | --- |
| Male | JP segment | **2002-2006** | **↓** | **2002-2005** | **↑** | **2002-2010** | **↓** | **2002-2005** | **↑** | **2002-2006** | **↑** |
|  | JP segment | 2006-2009 | → | **2005-2020** | **↓** | **2010-2022** | **↑** | 2005-2019 | → | 2006-2019 | → |
|  | JP segment | **2009-2023** | **↓** | **2020-2023** | **↓** |  |  | **2019-2023** | **↓** | **2019-2023** | **↓** |
|  | Bk segment | 2002-2004 | → | **2002-2004** | **↑** | **2002-2004** | **↓** | **2002-2004** | **↑** | 2002-2004 | → |
|  | Bk segment | 2004-2007 | → | **2004-2010** | **↓** | 2004-2007 | → | **2004-2012** | **↓** | 2004-2012 | → |
|  | Bk segment | 2007-2015 | → | **2010-2020** | **↓** | **2007-2013** | **↑** | **2012-2020** | **↓** | 2012-2020 | → |
|  | Bk segment | 2015-2023 | → | **2020-2023** | **↓** | **2013-2020** | **↑** | **2020-2023** | **↓** | 2020-2023 | → |
|  | Bk segment |  |  |  |  | **2020-2023** | **↑** |  |  |  |  |
| Female | JP segment | **2002-2023** | **↓** | **2002-2006** | **↑** | **2002-2008** | **↓** | **2002-2006** | **↑** | **2002-2008** | **↑** |
|  | JP segment |  |  | 2006-2019 | → | **2008-2023** | **↑** | **2006-2023** | **↓** | **2008-2023** | **↓** |
|  | JP segment |  |  | **2019-2023** | **↓** |  |  |  |  |  |  |
|  | Bk segment | 2002-2004 | → | 2002-2004 | → | 2002-2004 | → | 2002-2004 | → | 2002-2004 | → |
|  | Bk segment | 2015-2018 | → | 2004-2012 | → | 2004-2012 | → | 2004-2012 | → | 2004-2013 | → |
|  | Bk segment | 2018-2023 | → | 2012-2020 | → | 2012-2019 | → | 2012-2020 | → | **2013-2020** | **↓** |
|  | Bk segment |  |  | 2020-2023 | → | 2019-2023 | → | 2020-2023 | → | 2020-2023 | → |

JP = Joinpoint segments (derived from Joinpoint regression); Bk = Breakpoint segments (derived from OLS segmented regression using BIC and Chow test); ↑ = increasing trend; ↓ = decreasing trend; → = stable or non-significant trend; Time intervals correspond to detected segments within each model. Significant values are highlighted in bold.

**Figure S1: Trends in age-standardised death rates (SDRs) for suicide, undetermined intent, and unintentional deaths in Portugal by sex, 2002–2023**

|  | **males** | **females** |
| --- | --- | --- |
| **Suicide** | 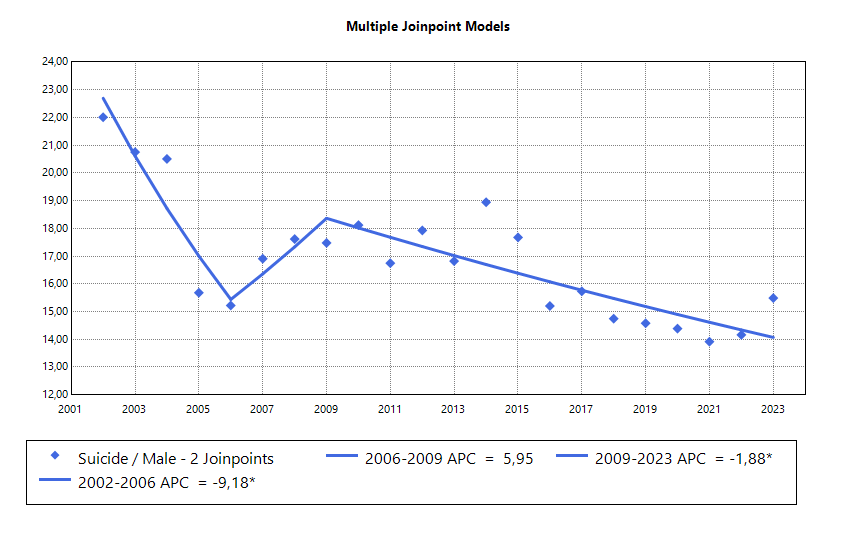 | 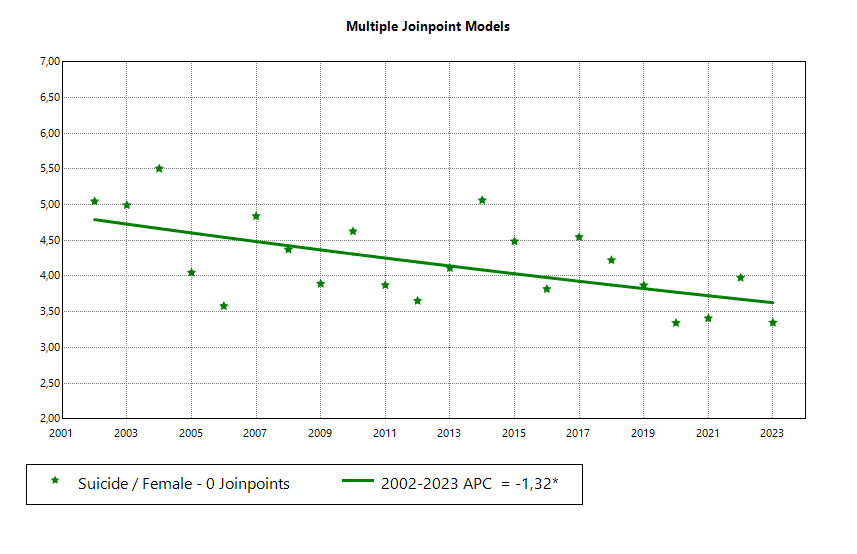 |
| **Undetermined** | 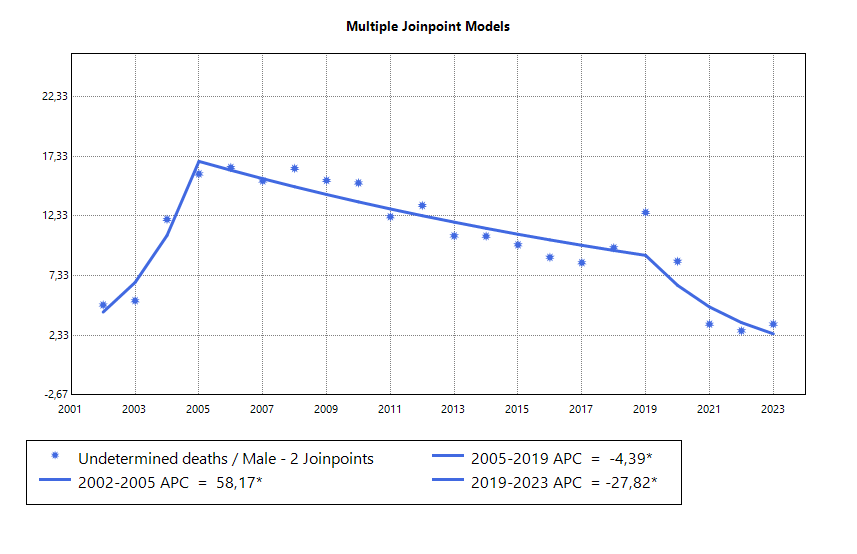 | 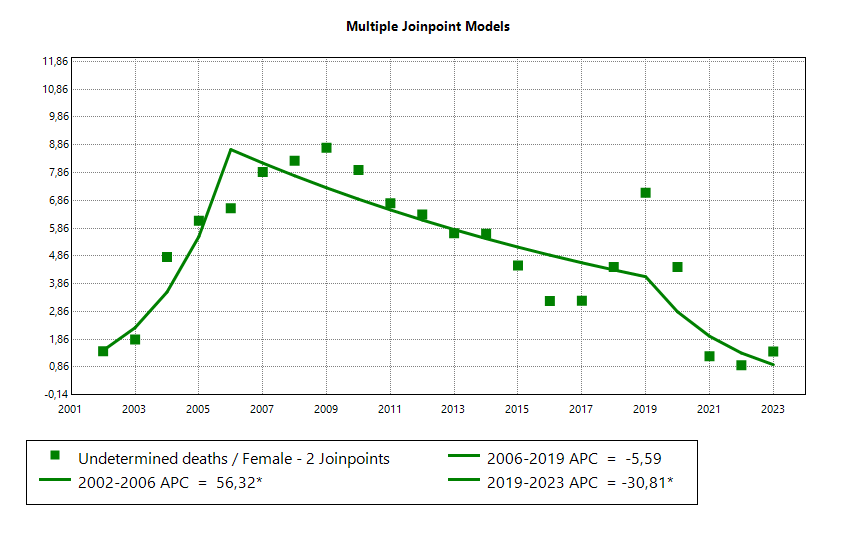 |
| **Unintentional** | 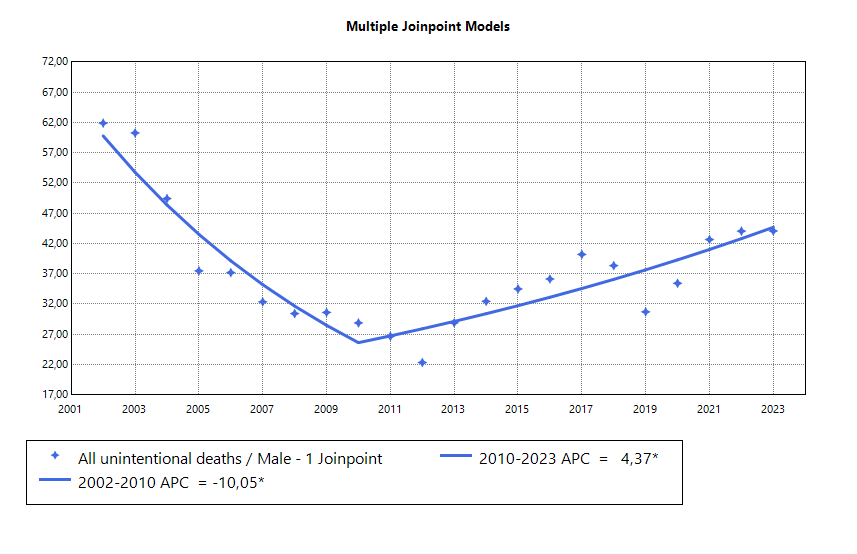 | 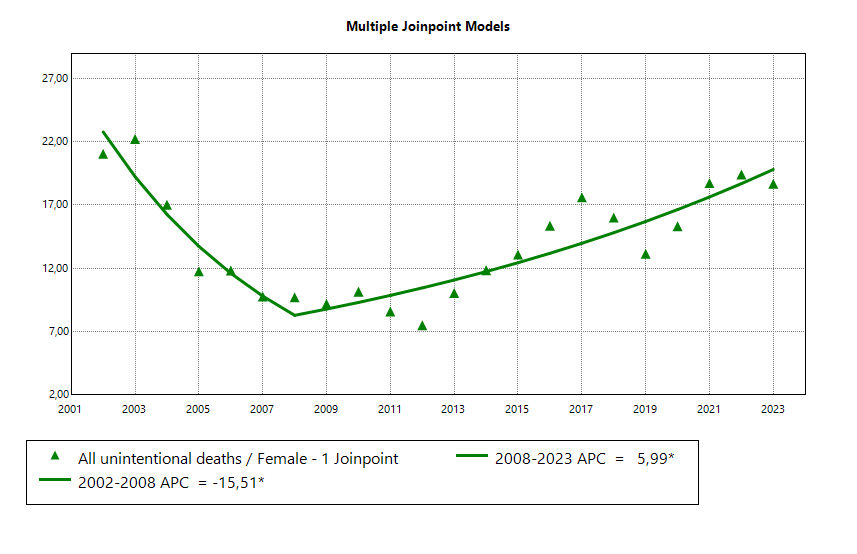 |

**Figure S2: Structural breakpoints in age-standardised death rates (SDRs) for undetermined intent-to-suicide ratio and for undetermined intent-to-unintentional deaths in Portugal by sex, 2002–2023**

|  | **2004** (2003–2005); **2010** (2008–2011); 2020 (2019–2021) | 2004 (2003–2005); 2012 (2008–2013); 2020 (2019-2022) |
| --- | --- | --- |
| **UnD : Suic** | 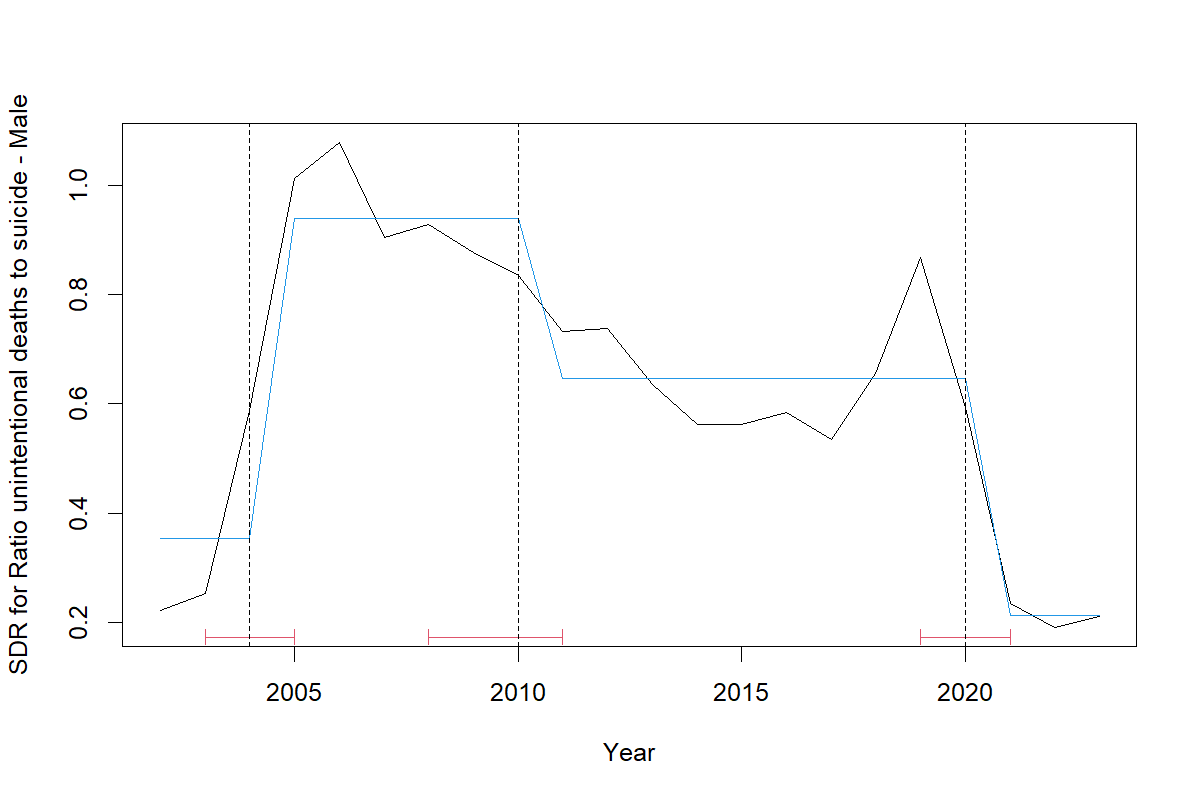 | 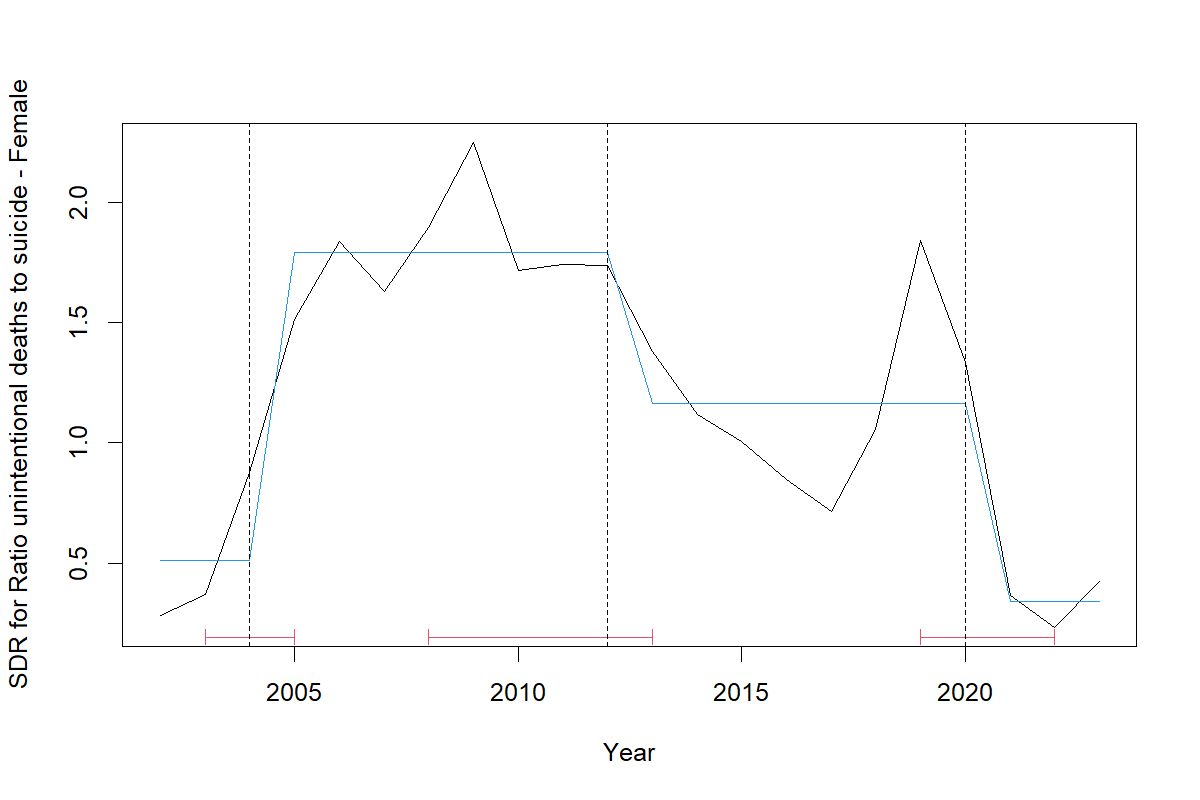 |
|  | 2004 (2003–2005); 2012 (2010–2013); 2020 (2020-2020) | 2004 (2003–2005); **2013** (2012–2015); 2020 (2020-2020) |
| **UnD : Accs** | 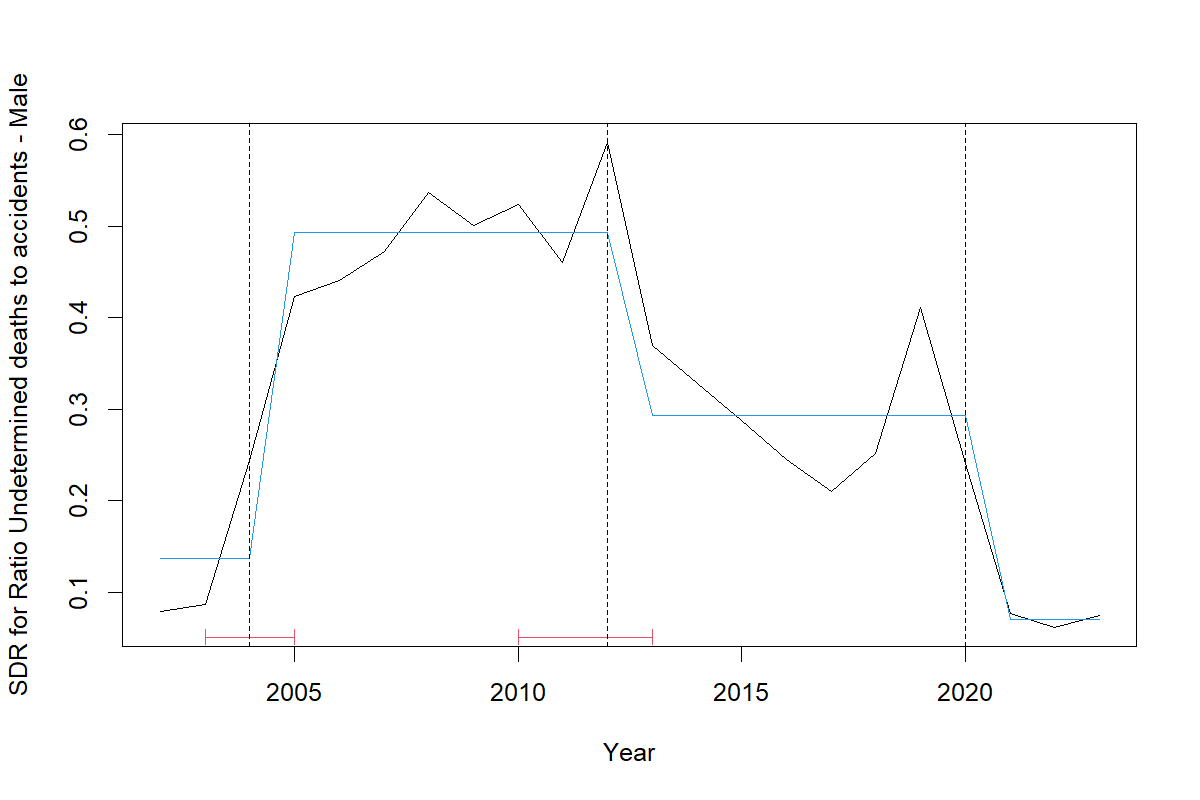 | 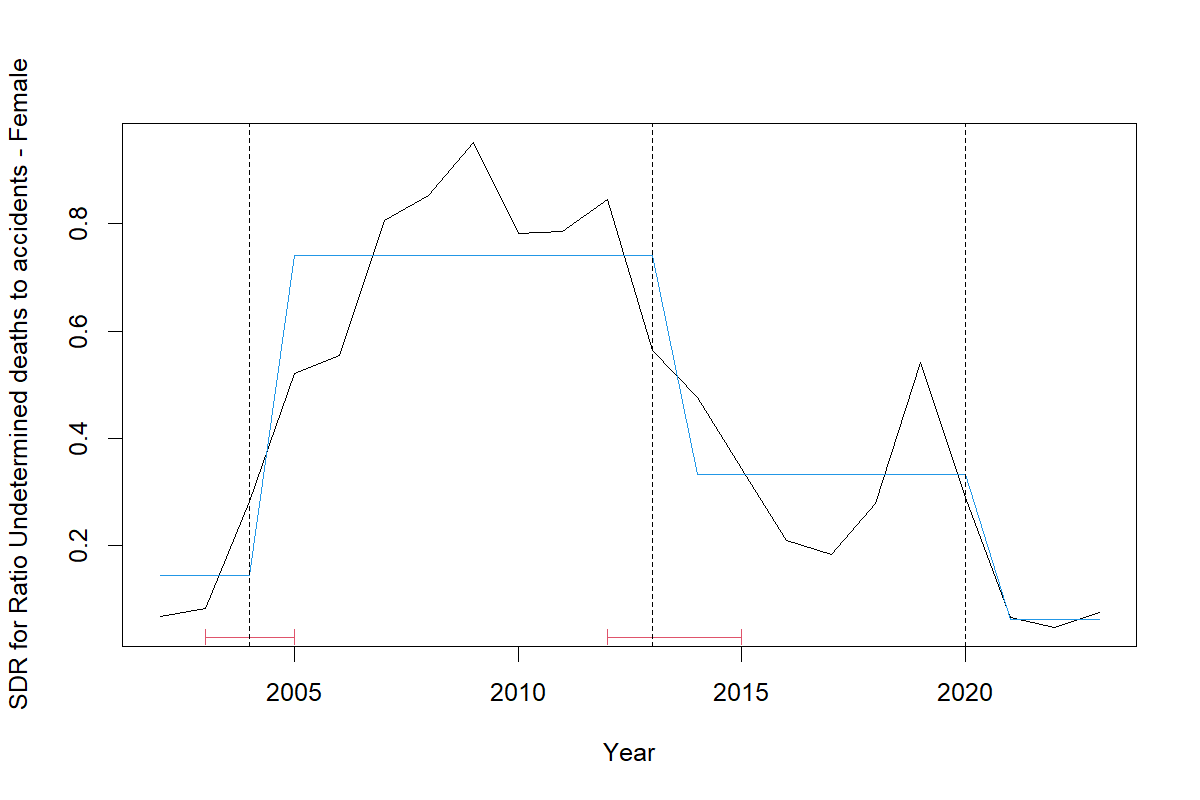 |

Note: Vertical dashed lines represent statistically identified breakpoints from segmented regression models based on BIC and Chow test (p<0.01). Blue lines indicate Ordinary Least Squares regression segments. Breakpoint years are shown above each plot with 95% confidence intervals in brackets.
